# Thoracic epidural versus paravertebral blockade for reducing chronic post-thoracotomy pain (TOPIC-2): an open-label, allocation-concealed, multicentre, randomised controlled trial

**DOI:** 10.1101/2025.08.07.25333201

**Authors:** Ben Shelley, Lee Middleton, Rebecca Boyles, Michael Gilbert, Andreas Goebel, Ira Goldsmith, Stephen Grant, Louise Jackson, Mishal Javed, Sajith Kumar, Nandor Marczin, Philip McCall, Rajnikant Mehta, Teresa Melody, Babu Naidu, Sridhar Rathinam, Hannah Summers, Lajos Szentgyorgyi, Sarah Tearne, Ben Watkins, Matthew Wilson, Andrew Worrall, Joyce Yeung, Fang Gao Smith, the TOPIC-2 Investigators

## Abstract

**Importance:** Many patients undergoing thoracotomy suffer from debilitating chronic post-thoracotomy pain (CPTP) lasting months or years postoperatively. The effectiveness of the commonly used two analgesic techniques, paravertebral blockade (PVB) and thoracic epidural blockade (TEB), on the incidence of CPTP is unknown.

**Objective:** To test the hypothesis that PVB reduces the incidence of CPTP compared with TEB.

**Design:** Pragmatic, open-label, allocation-concealed randomized controlled trial. Participants were recruited between January 8th, 2019 and September 29th, 2023.

**Setting:** 15 UK thoracic centers.

**Participants:** 770 eligible adult patients undergoing thoracotomy were randomly assigned (1:1) to TEB or PVB using a web-based randomization service.

**Intervention(s):** Participants in the PVB group received three single-shot paravertebral injections of local anesthetic before knife-to-skin, followed by placement of a paravertebral catheter. Participants in the TEB group had a thoracic epidural catheter placed and loaded with local anaesthetic before knife-to-skin.

**Main Outcome(s) and Measure(s):** The primary outcome was the incidence of CPTP at 6 months post-randomization, defined as a 100mm Visual Analogue Score (VAS) greater than or equal to 40mm (indicating moderate pain) when considering ‘worst chest pain over the last week’. Secondary outcomes included additional measures of chronic/acute pain, complications and quality of life.

**Results:** The trial enrolled 770 patients (342 female patients (44.4%); mean (SD) age, 66.6 (11.0) years). After 33 post-randomisation exclusions of patients who did not proceed to thoracotomy, 737 were included in the modified intention-to-treat population (364 PVB, 373 TEB). At 6-months, 59 (22%) of 272 participants in the PVB group and 47 (16%) of 292 participants in the TEB group developed CPTP (adjusted risk ratio=1·32 [95%CI 0·93 to 1·86]; adjusted risk difference=0·05 [95%CI –0·01 to 0·11]; p=0·12). During the acute phase, pain was greater on day 1 with PVB, but not different on days 2-3. Hypotension was less common in the PVB group; complications were similar otherwise.

**Conclusions and Relevance:** PVB did not reduce the incidence of CPTP at 6 months compared to TEB. TEB appeared to provide marginally better acute pain relief on postoperative day 1, but there was no difference thereafter. Postoperative complications were comparable between groups. The findings support the ongoing utility of both techniques.

**Trial Registration:** This trial is registered with ClinicalTrials.gov, NCT03677856.

**Funding Organisation:** NIHR HTA reference-16.111.111

**Key Points:** *Question:* Does paravertebral blockade (PVB) reduce the incidence of chronic post-thoracotomy pain (CPTP) compared to thoracic epidural blockade (TEB)?

*Findings:* In this open-label, allocation-concealed, multicenter randomised clinical trial that included 770 adults, paravertebral blockade did not significantly reduce the incidence of CPTP six months postoperatively compared to thoracic epidural blockade (22% with PVB vs 16% with TEB).

*Meaning:* In adult patients undergoing thoracotomy, providing acute perioperative analgesia with PVB does not reduce the incidence of CPTP compared to TEB.

## Introduction

Thoracotomy surgery is considered one of the most painful surgical procedures and can cause chronic post-surgical pain lasting months to years postoperatively. The presence of chronic post-thoracotomy pain (CPTP), defined as pain that recurs or persists at least three months following surgery^1^, has been reported to occur with an incidence as high as 50%^2^. CPTP can be severe and debilitating to patients, leading to wide-ranging impacts on functional activity and quality of life^3–5^. The well-recognised burden of chronic pain following surgery is such that this was identified as a top 10 research priority by the James Lind Alliance through the Anaesthesia and Perioperative Care Priority Setting Partnership^6^.

Aggressive management of acute pain resulting from thoracotomy may reduce the likelihood of developing chronic pain^7^. Two main analgesic techniques are commonly used for postoperative pain control following thoracotomy, thoracic epidural blockade (TEB) and paravertebral blockade (PVB), which both seek to block afferent nociceptive transmission at a spinal cord level preventing ascending transmission. Some suggest that by unilaterally blocking afferent nerve transmission at the paravertebral space, PVB may more effectively block nociceptive transmission than TEB^8,9^. Further, lying anteriorly in the PVB space, the sympathetic chain may be more effectively blocked by PVB than TEB^8^. In our pilot feasibility trial (TOPIC-1), the incidence of CPTP six-months postoperatively appeared lower with PVB but a definitive trial was required to confirm this finding reliably^10^.

Whilst there is good evidence that PVB provides similar analgesia to TEB for acute pain^7,11–15^, their comparative effects on chronic pain are unknown. A Cochrane review of 14 studies (698 participants) comparing the two techniques was forced to conclude that there was insufficient data on chronic pain to allow comparison for this endpoint^11^.

The TOPIC-2 trial was therefore conceived to test the hypothesis that in patients undergoing thoracotomy, the use of PVB for post-operative acute pain relief reduces the 6-month incidence of CPTP compared with TEB.

## Methods

### Study design

This was a multicenter, open-label, allocation-concealed, parallel group randomised controlled trial in 15 UK thoracic surgical centres. Ethics approval was obtained from the South-East Scotland Research Ethics Committee (REC 18/SS/0131). A trial steering committee provided independent oversight of the trial. Confidential inspection of available data was reviewed by a data monitoring committee annually; no reason to recommend halting or modifying the trial was identified. This trial is registered with ClinicalTrials.gov (NCT03677856), and the protocol has been published^16^.

### Participants

Eligible participants were adult patients (≥18) undergoing elective open thoracotomy who were willing to complete study follow-up questionnaires. Participants were excluded if they had a contraindication to TEB or PVB, a planned rib/chest wall resection or pleurectomy, had a previous thoracotomy on the same side or had undergone a median sternotomy within the past 90 days. All participants provided written informed consent.

### Randomization and masking

Participants were randomly assigned (1:1) to receive TEB or PVB through a secure online randomisation system, with the use of a minimisation algorithm for gender, age (<65 or ≥65 years), operation indication (lung cancer or other) and recruiting center. Due to the nature of intervention delivery, blind treating clinicians and research staff were considered impractical.

### Study interventions

Thoracic anaesthetists and surgeons performed trial interventions with confirmed experience in the techniques following review of training material. Local anaesthetic was delivered by continuous infusion through either an epidural or paravertebral catheter for up to 48 hours postoperatively. PVB delivery consisted of three single-shot injections, at an appropriate spinal level supplying the skin over the incision site, before the start of surgery. A catheter was then placed under direct vision during surgery, with a loading dose of local anaesthetic given before chest closure followed by continuous paravertebral infusion. For TEB, an epidural catheter was inserted at the spinal level supplying the skin at the incision site, followed by a test and loading dose of local anaesthetic before the start of surgery and an infusion for postoperative use.

Some variations in the technical aspects of intervention delivery were permitted. This pragmatic approach reflected minor variations in real-life practice as judged by the local team to be best for the individual patient. This included single-shot PVB insertion using ultrasound or landmark techniques, choice of local anaesthetic, and addition of opioids to local anaesthetic mixtures.

### Outcomes

Our primary outcome was the presence of chronic post-thoracotomy pain (CPTP) at 6 months post-randomisation. Participants were asked to indicate their ‘worst chest pain over the last week’ on a 100mm visual analogue scale (VAS; 0–100). The presence of CPTP was defined as VAS≥40mm, indicating at least moderate pain^17^.

Secondary outcomes consisted of: both worst and average chest pain VAS scores in the acute (days 1-3 and discharge) and chronic (3, 6 and 12 months) phases postoperatively, Brief Pain Inventory (BPI), Short Form McGill Pain Questionnaire 2 (SF-MPQ-2), health-related quality of life (EQ-5D-5L), hospital anxiety and depression score (HADS), patient satisfaction (Likert scale), analgesia use, and complications (analgesic, surgical and pulmonary), critical care admission, mortality and serious adverse events.

### Sample size

The sample size assumed a 30% incidence of CPTP in the TEB group, based on data observed in a systematic review^16^ and TOPIC-1^12^. To detect a plausible 10% absolute reduction in the PVB group with 90% power (two-sided α=0.05), 392 participants per group were required. Assuming a 10% death rate by 6 months and a 15% loss to follow-up (as per TOPIC-1), we aimed to recruit 1026 participants. In response to a slower-than-expected recruitment rate, in August 2023, the funder recommended that this target be revised to 770 participants, reducing power from 90% to 80%, but facilitating trial completion.

### Statistical analysis

A statistical analysis plan (appendix pp 33-57) was written and reviewed by the oversight committees before any analyses took place. Analyses were carried out in a modified intention-to-treat population (all participants randomised where a thoracotomy took place and were alive at the corresponding assessment timepoint). Estimates of differences between groups were presented with 95%, two-sided confidence intervals, adjusting for the minimisation variables (fixed effects apart from centre as random).

Repeated binary measures were analysed using generalised estimating equations to estimate risk difference (identity link) and relative risk (log link), incorporating all relevant timepoints (acute or chronic). A time-by-treatment interaction term was included to account for temporal effects. For the primary outcome, the number of observations included can be seen in Figure 1.

**Figure 1:**
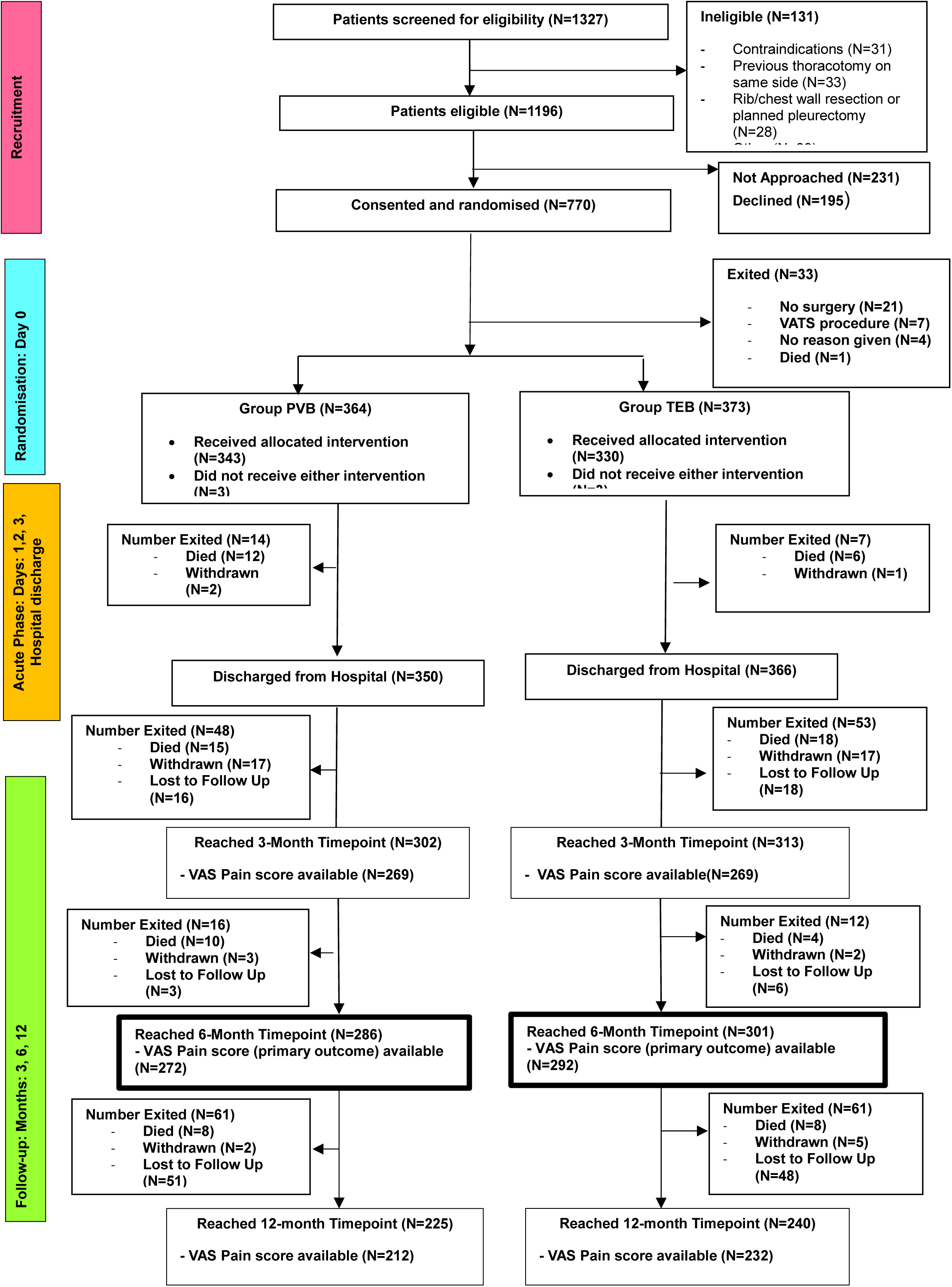
Trial profile

For secondary outcomes measured continuously, adjusted mean differences were produced from mixed linear regression models, incorporating all time points. Baseline values were included, where appropriate. Where the data was skewed, we presented median differences with confidence intervals from bootstrapping methods. Ordinal data were analysed using a generalised estimating equation model to estimate a common odds ratio and confidence intervals.

Sensitivity analyses on the primary outcome included a per-protocol analysis (participants who received their randomized intervention), an analysis considering sustained chronic pain (CPTP at 3 and 6 months), and a tipping point analysis to explore any potential impact of missing data. Subgroup analyses were prespecified and examined by including the subgroup by treatment interaction parameter into the model.

Analyses were performed in SAS (version 9.4) or Stata (version 18).

## Results

Between January 2019 and September 2023, 1327 participants were screened for eligibility, 770 were randomly assigned to PVB (n=386) or TEB (n=384) (Figure 1). Thirty-three patients did not undergo thoracotomy (21 of whom did not have any surgery and 7 received minimally invasive surgery due to perioperative changes to the surgical plan). Withdrawal and loss to follow-up rates were similar across groups. The follow-up rate for the primary outcome, excluding post-operative deaths (n=65) and those who did not undergo thoracotomy, was 564 (84%) of 672 participants.

At trial enrolment, baseline characteristics were balanced between groups (Table 1; appendix pp3-6 ST1-2). The mean age was 66.6 years (SD 11·0), 428 (56%) were male, and 658 (85%) were being treated for lung cancer. Reflecting the UK thoracic surgical population, the majority of patients were American Society of Anesthetists (ASA) class II or above, reported no, or mild functional limitation, had mild to moderate impairment of lung function and high rates of comorbidity (Table 1, appendix pp5 ST2). The predominant indication for thoracotomy was lobectomy or bilobectomy (498, 68%) (Table 1), which was performed via a posterolateral muscle-sparing approach in 704 (96%) of cases (appendix pp7 ST3). Importantly, aspects of surgical technique which are recognised to contribute to the risk of chronic pain development such as the rates of muscle and intercostal nerve-sparing surgery, rib resection and fracture, closure technique and the number of intercostal drains inserted, which were not protocolised, were well balanced between groups (appendix pp7 ST3).

**Table 1:** Baseline Characteristics and Operative Details.

| Baseline Details (in all randomized) |  |  |  |
| --- | --- | --- | --- |
|  |  | **PVB**<br>**(N=386)** | **TEB**<br>**(N=384)** |
| Gender | Male | 216 (56%) | 212 (55%) |
|  | Female | 170 (44%) | 172 (45%) |
| Age (years) | <65 years | 151 (39%) | 150 (39%) |
|  | ≥65 years | 235 (61%) | 234 (61%) |
|  | Mean (SD) | 66.5 (10.6) | 66.6 (11.4) |
| Indications for thoracotomy | Lung cancer | 330 (85%) | 328 (85%) |
|  | Other | 56 (15%) | 56 (15%) |
| BMI (kg/m<sup>2</sup>) | Mean (SD) | 27.6 (5.5) | 27.4 (5.4) |
|  | Missing | 2 | 3 |
| ECOG performance status | 0: Normal activity | 214 (59%) | 229 (63%) |
|  | 1: Symptomatic but nearly fully ambulatory | 141 (39%) | 130 (36%) |
|  | 2: Symptomatic but bed <50% daytime | 8 (2%) | 7 (2%) |
|  | 3: Symptomatic bed>50% daytime | 1 (<1%) | 0 (-) |
|  | 4: Unable to get out of bed | 0 (-) | 0 (-) |
|  | Not Assessed / Missing | 22 | 18 |
| ASA status assessed | I: Normal healthy | 15 (4%) | 13 (4%) |
|  | II: Mild systemic disease | 131 (37%) | 126 (36%) |
|  | III: Severe systemic disease | 199 (56%) | 195 (56%) |
|  | IV: Constant threat to life | 10 (3%) | 13 (4%) |
|  | V: Who is not expected to survive without the operation | 0 (-) | 0 (-) |
| FEV1 % predicted | Mean (SD) | 85.5 (19.4) | 85.0 (20.4) |
|  | Missing / Not Assessed | 33 | 30 |
| FEV1 / FVC | Mean (SD) | 0.68 (0.11) | 0.68 (0.11) |
|  | Missing / Not Assessed | 40 | 31 |
| DLCO % predicted | Mean (SD) | 78.5 (18.6) | 75.1 (19.1) |
|  | Missing / Not Assessed | 87 | 81 |
| Operative Details (for all who progressed to thoracotomy) |  |  |  |
|  |  | **PVB**<br>**(N=364)** | **TEB**<br>**(N=373)** |
| Duration of surgery (mins) | Median [IQR] | 167 [127 – 222] | 160 [116 – 206] |
|  | Missing | 6 | 12 |
| Primary operation type | Lobectomy/bilobectomy | 242 (66%) | 256 (69%) |
|  | Sub lobar Resection | 57 (16%) | 68 (18%) |
|  | Pneumonectomy | 27 (7%) | 21 (6%) |
|  | Diaphragmatic | 11 (3%) | 13 (4%) |
|  | Open & Close | 9 (2%) | 9 (2%) |
|  | Resection of mediastinal tumour/cyst | 3 (<1%) | 5 (1%) |
|  | Resection of airways without the removal of the lung | 2 (<1%) | 1 (<1%) |
|  | Other | 4 (1%) | 9 (2%) |
| Side of operation | Left | 161 (44%) | 169 (45%) |
|  | Right | 202 (55%) | 204 (55%) |
|  | Missing | 1 | 0 |
| Thoracotomy performed | Anterior | 12 (3%) | 19 (5%) |
|  | Posterolateral | 350 (97%) | 354 (95%) |
|  | Missing | 2 | 0 |

ASA = American Society of Anaesthesiologists, ECOG = Eastern Cooperative Oncology Group, FEV1 = Forced Expiratory Volume in 1 Second, FVC=Forced Vital Capacity, DLCO=Diffusing Capacity for Carbon Monoxide

Six hundred and seventy-three (91%) of 737 participants undergoing thoracotomy received the randomised allocation; the most common reason for non-adherence was technical difficulties/inability to perform the randomised intervention (n=31, 48%) (appendix pp8 ST4).

During the acute post-operative phase, rates of non-opioid and straightforward adjunctive analgesia use were not different between groups, whilst systemic (oral or intravenous) strong opioid administration was more common in the PVB group (appendix pp12-13 ST4).

Baseline VAS scores were low and balanced between groups (appendix pp 26 ST11). We found no statistically significant difference between groups in the primary outcome of CPTP. Fifty-nine (22%) of 272 participants in the PVB group and 47 (16%) of 292 participants in the TEB group reached this endpoint (adjusted RR 1·32 [95% CI 0·93 to 1·86]; adjusted RD 0·05 [95% CI –0·01 to 0·11]; p=0·12; Table 2; Figure 2). Supportive analyses, including a per-protocol analysis, had minimal impact on estimates (appendix pp19 ST9). For the tipping-point analysis, the rate of primary outcome in the PVB group in those with missing responses would need to be 25.5% or higher to materially impact the results (appendix pp20-21 SF1-2). There was no evidence of any differential effect in the subgroups (appendix pp18 ST8).

**Figure 2:**
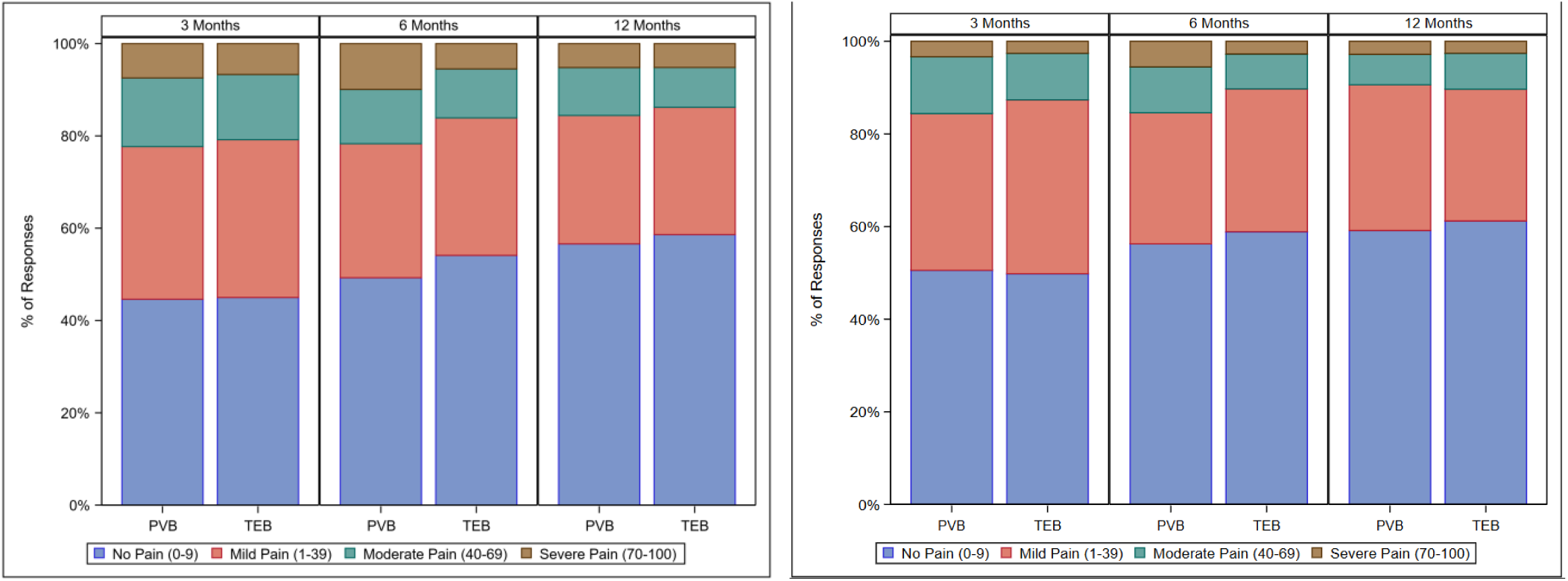
Worst (a) and average (b) chronic pain severity over time. By 100mm VAS categories. Data was presented for n=538, 564, and 444 respondents at 3, 6, and 12 months, respectively. See supplementary appendix table 7 for analysis.

**Table 2:** VAS ^1^ Chronic Pain Responses.

|  | **PVB**<br>N / N (%) | **TEB**<br>N / N (%) | Adjusted Risk<br>Difference<sup>2</sup> (95%<br>CI) | Adjusted Relative<br>Risk<sup>3</sup> (95% CI) |
| --- | --- | --- | --- | --- |
| **Primary Outcome: Worst Pain (with regards to chest) VAS ≥ 40 at 6 months** |  |  |  |  |
| 6 Month | 59 / 272 (22%) | 47 / 292 (16%) | 0.05 (-0.01, 0.11) | 1.32 (0.93, 1.86) |
| **Worst Pain (with regards to chest) VAS ≥ 40 at other timepoints** |  |  |  |  |
| 3 Month | 60 / 269 (22%) | 56 / 269 (21%) | 0.01 (-0.06, 0.07) | 1.08 (0.78, 1.49) |
| 12 Month | 33 / 212 (16%) | 32 / 232 (14%) | 0.02 (-0.05, 0.08) | 1.12 (0.72, 1.74) |
| **Average Pain (with regards to chest) VAS ≥ 40** |  |  |  |  |
| 3 Month | 42 / 269 (16%) | 34 / 269 (13%) | 0.02 (-0.04, 0.08) | 1.23 (0.81, 1.88) |
| 6 Month | 42 / 272 (15%) | 30 / 292 (10%) | 0.04 (-0.01, 0.10) | 1.48 (0.95, 2.30) |
| 12 Month | 20 / 213 (9%) | 24 / 232 (10%) | 0.00 (-0.05, 0.05) | 0.90 (0.51, 1.56) |

<sup>1</sup> VAS scored from 0 to 100 with 100 indicating the worst pain imaginable. The presence of at least moderate pain is indicated by scores greater than or equal to 40mm.
<sup>2</sup> Adjusted for minimisation variables. Estimated Risk Difference (RD) <0 indicates less pain with PVB.
<sup>3</sup> Adjusted for minimisation variables. Relative Risk (RR) <1 favours the PVB group.

During the chronic phase, most of the patient-reported pain and quality of life secondary outcomes favoured TEB on average. Still, differences were generally minor, not statistically significant and inconsistent over the time frame observed (appendix pp22 ST10). Chronic phase use of conventional, opioid and neuropathic analgesics was similar at all time-points postoperatively (appendix pp28 ST12). During the acute phase, both worst and average pain were higher on day 1 post-operatively with PVB (adjusted mean difference in VAS score: 7.7mm [95% CI 2.8-12.5] and 7.0mm [95% CI 2.7-11.2] respectively; appendix pp26 ST11). There were no differences in worst or average pain VAS, nor BPI on postoperative days 2-3.

Hypotension (defined as systolic blood pressure <90mmHg) (RR 0.66 [95% CI 0.46–0.94]) and pruritis (RR 0.37 [95% CI 0.21–0.64]) were less common in the PVB group, whilst more participants experienced urinary retention in the PVB group (RR 2.21 [95% CI 1.35–3.61], Table 3). Similar levels of surgical and postoperative pulmonary complications were observed in each group. There was no difference between groups in critical care and hospital length of stay or in-hospital mortality (Table 3). Forty-three participants (12%) in the PVB group and 33 (9%) in the TEB group experienced SAEs; only one in each group was considered related to the allocated intervention.

**Table 3:**
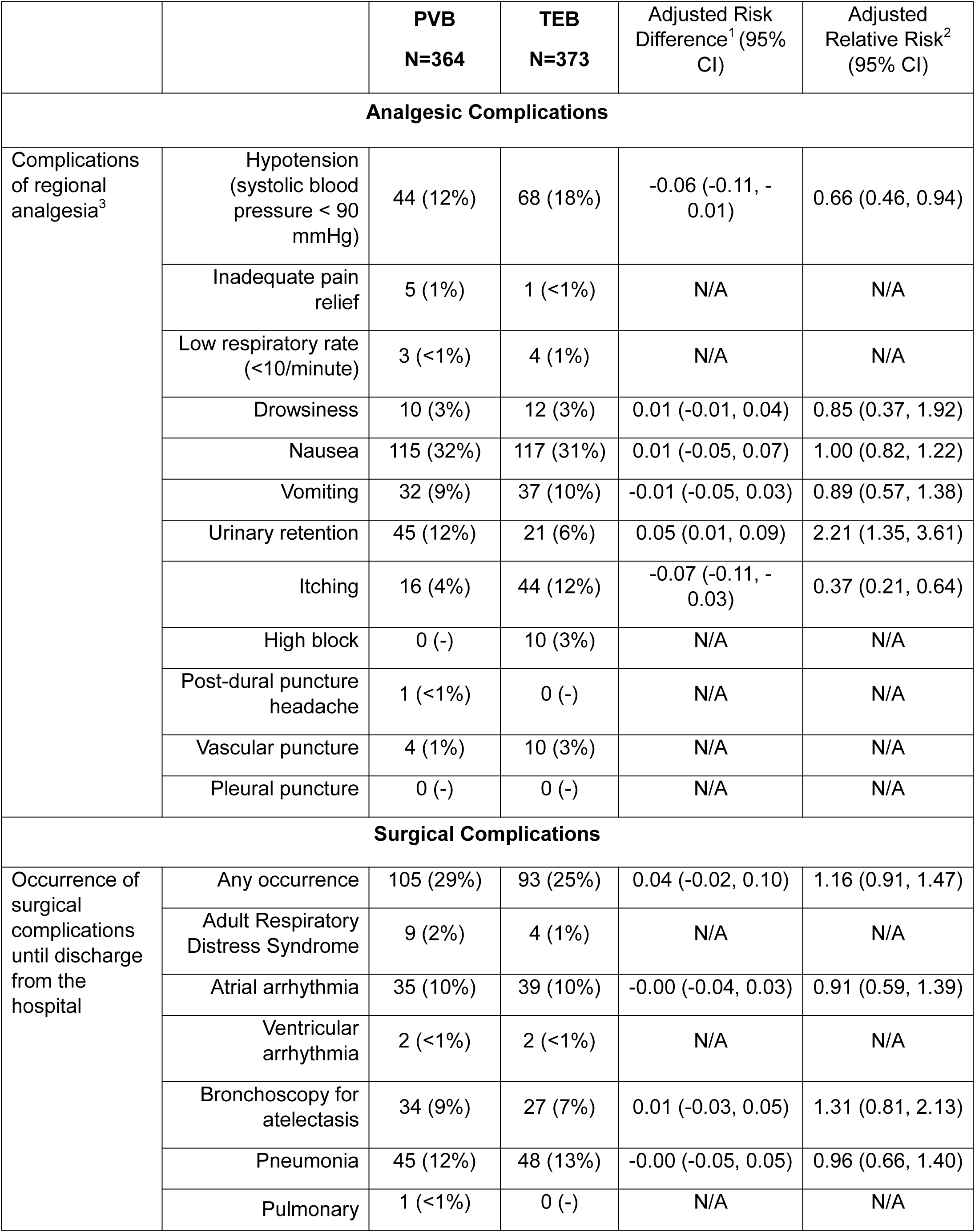
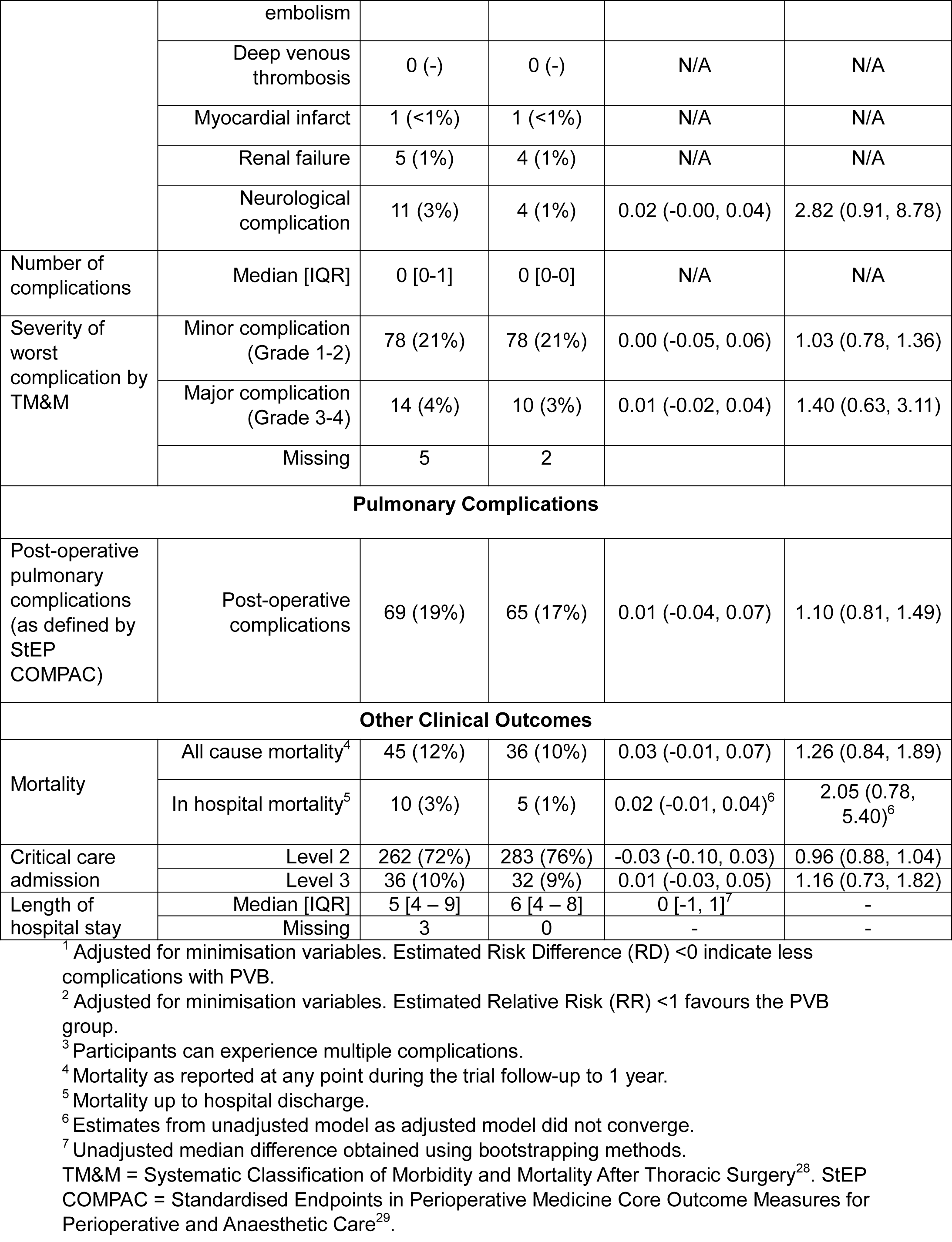
Analgesic, surgical and postoperative pulmonary complications and length of stay.

## Discussion

In this pragmatic, multicentre, randomised trial, we found no evidence of any difference in the incidence of CPTP between patients receiving TEB versus PVB. CPTP, defined according to our primary outcome definition, occurred more commonly in patients receiving PVB (22% vs 16%), but this difference was not statistically significant. Across the study as whole, the “signal” of no difference between techniques is compelling, being further supported by no differences in a large volume of secondary endpoint data including CPTP defined by ‘average’ rather than ‘worst’ pain VAS, Brief Pain Inventory and SF-MPQ-2 Inventory scores and the observation of similar analgesic use between groups over twelve months of follow-up. Whilst no consistent differences in outcomes were observed, our primary outcome result strongly suggests that our original hypothesis of PVB being superior to TEB in preventing CPTP is unlikely to be true, given the confidence interval around the treatment effect estimate excludes a clinically meaningful difference in favour of PVB.

In addition to several inconclusive observational studies, we know four other randomized controlled trials (including TOPIC-1) of TEB vs PVB, which examined chronic pain outcomes. All were underpowered for a comparison of chronic pain incidence (sample sizes ranged from 50-100) and either found no difference between techniques^18^ or a modest reduction in chronic pain in the PVB^10,19^ or TEB^20^ groups, which was not statistically significant. Though our results now allay this uncertainty by demonstrating that PVB, when compared to TEB, is not any more effective in reducing the incidence of CPTP, our study further highlights the striking burden of chronic post-surgical pain following thoracic surgery. Across the study population as a whole, 19% of patients report moderate to severe CPTP 6-months postoperatively, a figure in keeping with previous reports^3^, but slightly less than we observed in TOPIC-1 (although within the 95%CI of 18%-42%)^10^. The fact that the observed rates were slightly lower than assumed in our sample size calculation may be inconsequential given that the results did not support the hypothesis of superiority with PVB.

Multiple systematic reviews and meta-analyses have concluded that PVB and TEB provide equally adequate analgesia in the acute postoperative phase^11–15^. Although we demonstrated a reduced ‘worst pain’ VAS on postoperative day 1 in the TEB group, the magnitude of this difference was slight, reflecting a mean difference (95%CI) of 7.7 (2.8,12.5) mm. Although matched by a similar reduction in ‘average pain’ VAS on postoperative day 1, there was no difference between groups on days 2 and 3, nor on hospital discharge. Further this difference on POD1 falls short of the accepted 10mm minimum clinically important difference^21^ on a 100mm VAS, further suggesting no clinically significant difference between groups.

The provision of excellent acute pain control is of paramount importance not just in avoiding patient distress and minimising the risk of subsequent chronic pain development, but also in facilitating engagement with physiotherapy and early mobilisation following surgery and reducing the risk of postoperative complications, particularly pulmonary complications. For this reason, as secondary endpoints, we built a robust postoperative complications dataset into the TOPIC-2 trial. In keeping with previous reports, minor complications of pruritis^11^ and hypotension^11–15^ occurred more commonly in the TEB group. Urinary retention in the current study was however more common in patients receiving PVB, a finding at odds with previous reports, but which is likely confounded by the high rates of urinary catheter placement in the TEB group (33% in the PVB group vs 72% in the TEB group (appendix pp8 ST4)). Overall, the incidence of both minor, and major postoperative complications was no different between the two groups. In-hospital and all-cause mortality was more common in the PVB group, but this result was not statistically significant and could easily be explained by chance.

Since the conception of the TOPIC-2 trial (the TOPIC-1 pilot study began recruitment in July 2015^10^), there have been significant advances in minimally invasive thoracic surgery with rapid increases in the rate of video- and robotically-assisted thoracoscopic surgery (VATS / RATS). Despite these advances, evidence suggests there remains a large and consistent population undergoing thoracotomy for whom the results of the TOPIC-2 trial remain important. Contemporary data from the UK Society of Cardiothoracic Surgeons suggest a stable thoracotomy population in excess of 1000 patients/year for lung cancer resection alone^22^. Data from the US Society of Thoracic Surgeons suggests a similar stabilisation in thoracotomy rate in recent years^23^. Rising VATS/RATS rates have been paralleled by an increase in overall resection rate^23^. This increase in overall resection rate further reflects developing practice to perform resection in patients with increasingly advanced disease^24^. Improved tumour shrinkage rates with neoadjuvant chemoimmunotherapy are redefining resectability criteria, but in many cases residual disease will necessitate thoracotomy^25^.

A significant strength of our study lies in its sample size; TOPIC-2 is by some margin the largest clinical trial comparing TEB and PVB conducted in the thoracic surgical population to date and recruited more patients than the 14 studies reported in the 2016 Cochrane review recruited cumulatively^11^. By conducting the trial in 15 centres, in all four nations across the UK, we provide a widely generalisable result. Protocol adherence within the trial appeared strong in both groups with low crossover rates, and compares favourably with other large randomised controlled trials of perioperative epidural blockade^26^.

Following the cessation of trial recruitment during the first wave of the COVID-19 pandemic, the research and development landscape in the UK was such that recruitment slowed significantly, and it became apparent that achieving our target sample size was not feasible. For this reason, in consultation with the funder, and with the support of the Trial Steering Committee and Independent Data Safety Monitoring Committee, but blind to the accumulated study data we elected to reduce the target sample size (from 1026 to 770) reflecting a reduction in statistical power from 90% to 80%. The impact of this reduction in sample size was ultimately immaterial given the findings.

The TOPIC-2 Trial was an open-label trial as we deemed it impractical to blind participants completely to the study intervention, and as such, a lack of blinding could be considered a limitation of our study. However, We do not believe this lack of blinding will have introduced significant bias to our results. Firstly, primary outcome data was collected via questionnaires administered by post, 6 months from the original operative procedure and are likely to be resilient to the effects of imperfect concealment. Secondly, participants were not explicitly informed of their treatment allocation; in our preceding pilot feasibility study, most participants were unaware of their treatment allocation at follow-up^12^. Thirdly, we have no reason to suspect that patients would have preconceived ideas regarding the relative efficacy of either intervention.

TOPIC-2 was conceived as a pragmatic clinical trial reflecting the ‘real-world’ variation in clinical practice across the NHS. Clinicians were therefore requested to perform the study interventions as practised in their institution. In the interests of achieving some degree of pre-emptive analgesia in both groups, we did, however, mandate the provision of three single-shot PVB injections before knife-to-skin in the PVB group. TEB catheters are commonly ‘loaded’ with local anaesthetic immediately following insertion, providing functional analgesia before surgical incision. The timing of placement of PVB is more variable, however, and the impact of block timing on analgesic efficacy is not fully understood^27^. It is conceivable that placing a PVB after knife-to-skin or indeed later in the surgical procedure could allow afferent nociceptive transmission implicated in the development of chronic pain to propagate before block placement. As such, the signal of no difference between PVB and TEB demonstrated by the TOPIC-2 trial can only be presumed in situations where intraoperative PVB catheter placement is preceded by preoperative blockade.

## Conclusion

Paravertebral blockade did not reduce the incidence of CPTP compared to TEB. In the acute phase both techniques provide adequate analgesia. TEB was associated with an increased incidence of hypotension and pruritis, but overall, the incidence of both minor and major postoperative complications was not different between groups.

## Contributors

FGS conceived the idea for this study and secured funding. LM led study design and protocol writing with assistance from BS, AG, SG, LJ, NM RM, LS, MW, AW, JY and FGS. JY led the literature search and systematic review with assistance from BN, MW and FGS. Study conduct and data collection were led by BS and FGS, with contributions from RB, MG, IG, SG, LJ, SK, NM, PM, RM, TM, BN, SR, LS, ST, BW, and JY. SG and AW are patient coinvestigators. TM led patient and public involvement in the trial. Study analysis and figure generation was done by HS, supervised by LM. All authors were involved in data interpretation. Writing of the paper was led by BS, LM, HS and FGS, with assistance from MG, AG, NM, PM, BN, SR, MW and JY.

## Data Sharing

The datasets generated during the current study will be made available by the Chief Investigator (FGS) upon reasonable request and in accordance with the Birmingham Clinical Trials Unit’s research collaboration and data transfer guidelines.

## Data Availability

The datasets generated during the current study will be made available by the Chief Investigator upon reasonable request and in accordance with the Birmingham Clinical Trials Unit's research collaboration and data transfer guidelines.

## Acknowledgments

The TOPIC-2 investigators would like to thank all the TOPIC-2 participants, their family members, nursing and medical staff, and the West Midlands Clinical Research Ambassador Group (patient and public involvement in research) (CRAG).

