## Supplementary Material for "Thoracic epidural versus paravertebral blockade for reducing chronic post-thoracotomy pain (TOPIC-2): an open-label, allocation-concealed, multicentre, randomised controlled trial"

**Title:**

### Supplementary Material

**Authorship:**

Ben Shelley*, Lee Middleton*, Rebecca Boyles, Michael Gilbert, Andreas Goebel, Ira Goldsmith, Stephen Grant, Louise Jackson, Nandor Marczin, Philip McCall, Rajnikant Mehta, Teresa Melody, Babu Naidu, Sridhar Rathinam, Hannah Summers, Lajos Szentgyorgyi, Sarah Tearne, Ben Watkins, Matthew Wilson, Andrew Worrall, Joyce Yeung, Fang Gao Smith, on behalf of the TOPIC-2 Investigators.

*Joint first authors.

### Table of Figures

Supplementary Figure 1 Tipping Point Scenario A........ 20

Supplementary Figure 2 Tipping Point Scenario B........ 21

**Supplementary Table 1: Other Baseline Characteristics**

| | | PVB (N=386) | TEB (N=384) |
|---|---|---|---|
| Centre | Glenfield Hospital | 85 (12%) | 84 (11%) |
| | Morriston Hospital Swansea | 60 (8%) | 62 (8%) |
| | Golden Jubilee National Hospital | 50 (7%) | 51 (7%) |
| | Queen Elizabeth Birmingham Hospital | 37 (5%) | 38 (5%) |
| | Castle Hill Hospital | 27 (4%) | 28 (4%) |
| | Harefield Hospital | 25 (3%) | 25 (3%) |
| | Wythenshawe Hospital | 24 (3%) | 26 (4%) |
| | Heartlands Hospital | 18 (2%) | 20 (3%) |
| | Essex Cardiothoracic Centre | 13 (2%) | 11 (1%) |
| | Nottingham City Hospital | 8 (1%) | 10 (1%) |
| | Belfast Royal Victoria Hospital | 6 (<1%) | 6 (<1%) |
| | Aberdeen Royal Infirmary | 3 (<1%) | 4 (<1%) |
| | Blackpool Victoria Hospital | 4 (<1%) | 3 (<1%) |
| | University Hospital Coventry | 2 (<1%) | 3 (<1%) |
| | John Radcliffe Hospital | 2 (<1%) | 2 (<1%) |
| Medical History[1] | Hypertension | 152 (39%) | 142 (37%) |
| | Cancer (not including current lung cancer) | 95 (25%) | 98 (26%) |
| | COPD | 90 (23%) | 82 (21%) |
| | Diabetes | 53 (14%) | 56 (15%) |
| | Ischaemic heart disease | 34 (9%) | 29 (8%) |
| | Hypothyroidism | 25 (6%) | 27 (7%) |
| | Previous stroke | 11 (3%) | 15 (4%) |
| | Hyperthyroidism | 10 (3%) | 10 (3%) |
| | Renal failure | 5 (1%) | 7 (2%) |
| | Congestive cardiac failure | 2 (<1%) | 3 (<1%) |
| Smoking Status | Never | 68 (18%) | 60 (16%) |
| | Current smoker | 44 (12%) | 42 (12%) |
| | Stopped < 6 weeks ago | 33 (9%) | 37 (10%) |
| | Stopped ≥6 weeks to <1 year ago | 55 (15%) | 50 (14%) |
| | Stopped ≥1 year ago | 171 (46%) | 175 (48%) |
| | Missing / Not Assessed | 15 | 20 |
| Smoking pack years[2] | Median [IQR], n | 34.5 [20 - 47], 282 | 34 [18 - 50], 288 |
| | Missing / Not Assessed | 36 | 36 |
| Alcohol consumption per week (units) | Median [IQR] | 1 [0 – 8] | 1 [0 – 8] |
| | Missing / Not Assessed | 39 | 50 |
| FEV1 | Mean (SD) | 2.28 (0.65) | 2.32 (0.74) |
| | Missing / Not Assessed | 35 | 29 |
| FVC | Mean (SD) | 3.36 (0.90) | 3.44 (1.00) |
| | Missing / Not Assessed | 39 | 31 |

| | | | |
|---|---|---|---|
| FVC % predicted | Mean (SD) | 100.0 (19.1) | 99.6 (20.3) |
| | Missing / Not Assessed | 37 | 35 |
| DLCO | Mean (SD) | 6.44 (2.10) | 6.60 (3.43) |
| | Missing / Not Assessed | 128 | 116 |

[1] Participants may have multiple other conditions

[2] Excluding those that have never smoked

**Supplementary Table 2: Baseline Characteristics in those who underwent thoracotomy (Modified Intention to Treat Population)**

| | | PVB<br>(N = 364) | TEB<br>(N= 373) |
|---|---|---|---|
| Gender | Male | 203 (56%) | 208 (56%) |
| | Female | 161 (44%) | 265 (44%) |
| Age | <65 years | 142 (39%) | 144 (39%) |
| | ≥65 years | 222 (61%) | 229 (61%) |
| | Mean (SD) | 66.5 (10.5) | 66.7 (11.5) |
| Thoracotomy: | Lung cancer | 311 (85%) | 319 (86%) |
| | Other | 53 (15%) | 54 (14%) |
| Centre | Glenfield Hospital | 85 (24%) | 84 (23%) |
| | Morriston Hospital Swansea | 60 (16%) | 62 (17%) |
| | Golden Jubilee National Hospital | 50 (14%) | 51 (14%) |
| | Queen Elizabeth Birmingham Hospital | 37 (10%) | 38 (10%) |
| | Castle Hill Hospital | 27 (7%) | 28 (8%) |
| | Harefield Hospital | 25 (7%) | 25 (7%) |
| | Wythenshawe Hospital | 24 (7%) | 26 (7%) |
| | Heartlands Hospital | 18 (5%) | 20 (5%) |
| | Essex Cardiothoracic Centre | 13 (4%) | 11 (3%) |
| | Nottingham City Hospital | 8 (2%) | 10 (3%) |
| | Belfast Royal Victoria Hospital | 6 (2%) | 6 (2%) |
| | Aberdeen Royal Infirmary | 3 (<1%) | 4 (1%) |
| | Blackpool Victoria Hospital | 4 (1%) | 3 (<1%) |
| | University Hospital Coventry | 2 (<1%) | 3 (<1%) |
| BMI (kg/m$^2$) | Mean (SD) | 27.6 (5.5) | 27.4 (5.2) |
| | Not Assessed / Missing | 0 | 1 |
| ECOG[1] performance status | 0: Normal activity | 203 (59%) | 224 (63%) |
| | 1: Symptomatic but nearly fully ambulatory | 132 (38%) | 127 (35%) |
| | 2: Symptomatic but bed <50% daytime | 7 (2%) | 7 (2%) |
| | 3: Symptomatic bed>50% daytime | 1 (<1%) | 0 (-) |
| | 4: Unable to get out of bed | 0 (-) | 0 (-) |
| | Not Assessed / Missing | 21 | 15 |
| ASA[2] status assessed | I: Normal healthy | 15 (4%) | 13 (4%) |
| | II: Mild systemic disease | 124 (37%) | 122 (36%) |
| | III: Severe systemic disease | 188 (56%) | 193 (57%) |
| | IV: Constant threat to life | 10 (3%) | 13 (4%) |
| | V: Who is not expected to survive without the operation | 0 (-) | 0 (-) |
| | Not Assessed / Missing | 27 | 32 |
| Medical History[3] | Hypertension | 144 (40%) | 140 (38%) |
| | Cancer (not including current lung cancer) | 93 (26%) | 98 (26%) |
| | COPD | 84 (23%) | 82 (22%) |

| | | | |
|---|---|---|---|
| | Diabetes | 51 (14%) | 55 (15%) |
| | Ischaemic heart disease | 32 (8%) | 28 (8%) |
| | Hypothyroidism | 24 (7%) | 27 (7%) |
| | Previous stroke | 11 (3%) | 15 (4%) |
| | Hyperthyroidism | 9 (2%) | 10 (3%) |
| | Renal failure | 4 (1%) | 7 (2%) |
| | Congestive cardiac failure | 2 (<1%) | 3 (<1%) |
| Smoking Status | Never | 67 (19%) | 58 (16%) |
| | Current smoker | 40 (11%) | 41 (11%) |
| | Stopped < 6 weeks ago | 32 (9%) | 37 (10%) |
| | Stopped ≥6 weeks to <1 year ago | 50 (14%) | 48 (13%) |
| | Stopped ≥1 year ago | 161 (46%) | 173 (48%) |
| | Missing / Not Assessed | 14 | 16 |
| Smoking pack years[4] | Median [IQR], n | 34.5 [20 – 49], 262 | 34 [18 – 50], 283 |
| | Missing / Not Assessed | 35 | 32 |
| Alcohol consumption per week (units) | Median [IQR] | 1 [0 – 8] | 1 [0 – 8] |
| | Missing / Not Assessed | 35 | 45 |
| FEV1 | Mean (SD) | 2.29 (0.65) | 2.33 (0.75) |
| | Missing / Not Assessed | 33 | 26 |
| FEV1 % predicted | Mean (SD) | 85.7 (19.4) | 85.1 (20.4) |
| | Missing / Not Assessed | 31 | 27 |
| FVC | Mean (SD) | 3.39 (0.91) | 3.45 (1.00) |
| | Missing / Not Assessed | 37 | 28 |
| FVC % predicted | Mean (SD) | 100.4 (19.2) | 99.7 (20.1) |
| | Missing / Not Assessed | 35 | 32 |
| FEV1 / FVC | Mean (SD) | 0.68 (0.11) | 0.68 (0.11) |
| | Missing / Not Assessed | 38 | 28 |
| DLCO | Mean (SD) | 6.45 (2.13) | 6.50 (3.34) |
| | Missing / Not Assessed | 117 | 110 |
| DLCO % predicted | Mean (SD) | 78.6 (18.8) | 75.2 (19.3) |
| | Missing / Not Assessed | 78 | 75 |

[1] Eastern Cooperative Oncology Group

[2] American Society of Anaesthesiologists

[3] Participants may have multiple other conditions

[4] Excluding those that have never smoked

**Supplementary Table 3: Additional Operative Details**

| | | **PVB (N=364)** | **TEB (N=373)** |
|---|---|---|---|
| Muscle sparing approach used | Yes | 289 (81%) | 286 (78%) |
| | No | 67 (19%) | 83 (22%) |
| | Missing | 8 | 4 |
| Serratus Muscles Spared | Yes | 280 / 288 (97%) | 274 / 282 (97%) |
| | No | 8 / 288 (3%) | 8 / 282 (3%) |
| | Missing | 1 | 4 |
| Latissimus Muscles Spared | Yes | 67 / 288 (23%) | 64 / 280 (23%) |
| | No | 221 / 288 (77%) | 216 / 280 (77%) |
| | Missing | 1 | 6 |
| Nerve spared | Yes | 231 (66%) | 232 (64%) |
| | No | 117 (34%) | 130 (36%) |
| | Missing | 16 | 11 |
| Rib-resection | Yes | 92 (26%) | 76 (21%) |
| | No | 266 (74%) | 290 (79%) |
| | Missing | 6 | 7 |
| Ribs fractured | Yes | 54 (15%) | 43 (12%) |
| | No | 306 (85%) | 326 (88%) |
| | Missing | 4 | 4 |
| Number of chest drains inserted | None | 3 (<1%) | 1 (<1%) |
| | 1 | 321 (89%) | 335 (90%) |
| | 2 | 39 (11%) | 37 (10%) |
| | Missing | 1 | 0 |
| Closure technique | Pericostal | 240 (67%) | 252 (69%) |
| | Rib punch and closure | 116 (33%) | 114 (31%) |
| | Missing | 8 | 7 |

**Supplementary Table 4: Intervention and Analgesia Details**

| | | PVB N = 364 | TEB N= 373 |
|---|---|---|---|
| **Protocol Adherence – Initial Block** | | | |
| Received the allocated intervention | | 343 (94%) | 330 (88%) |
| Received the other intervention (PVB or TEB) | | 18 (5%) | 40 (11%) |
| Received a different regional technique | | 3 (<1%) | 3 (<1%) |
| **Reasons for Non - Adherence** | | | |
| Technical difficulties | | 4 / 21 (19%) | 27 / 43 (63%) |
| Clinical Choice | | 10 / 21 (48%) | 7 / 43 (16%) |
| Patient Choice | | 1 / 21 (5%) | 4 / 43 (9%) |
| Other | | 2 / 21 (10%) | 1 / 43 (2%) |
| Unknown | | 4 / 21 (19%) | 4 / 43 (9%) |
| **Analgesic Intervention** | | | |
| Was local anaesthetic delivered pre-knife to skin? | Yes | 296 (96%) | 291 (96%) |
| | No | 11 (4%) | 13 (4%) |
| | Missing | 57 | 69 |
| Intraoperative monitoring | | | |
| Central line | Yes | 83 (23%) | 102 (28%) |
| | No | 271 (77%) | 259 (72%) |
| | Missing | 10 | 12 |
| Urinary catheter | Yes | 118 (33%) | 259 (72%) |
| | No | 236 (67%) | 103 (28%) |
| | Missing | 10 | 11 |
| Arterial line | Yes | 352 (99%) | 358 (99%) |
| | No | 4 (1%) | 5 (1%) |
| | Missing | 8 | 10 |
| Analgesic Intervention | | | |
| Block re-sited during surgery | No | 360 (99%) | 357 (96%) |
| | TEB | 2 (<1%) | 9 (2%) |
| | PVB | 2 (<1%) | 2 (<1%) |
| | Other | 0 (-) | 5 (1%) |
| Local Analgesia: TEB | | | |
| Level of Insertion | Median [IQR] | - | 5 [4 – 6] |
| | Missing | - | 1 |
| Opioid added to maintenance infusion | Yes | - | 283 / 317 (89%) |
| | Missing | - | 13 |
| Loading Bolus | | | |
| Bupivacaine | Used? | - | 40 / 324 (12%) |
| | Concentration (%), median [IQR] | - | 0.25 [0.163 – 0.25] |

| | | | |
|---|---|---|---|
| | Total volume (ml), median [IQR] | - | 7.75 [5 – 10] |
| Levobupivacaine | Used? | - | 283 / 324 (87%) |
| | Concentration (%), median [IQR] | - | 0.25 [ 0.25 – 0.25] |
| | Total volume (ml), median [IQR] | - | 10 [ 8 – 15] |
| Ropivacaine | Used? | - | 1 / 324 (<1%) |
| | Concentration (%), median [IQR] | - | 0.375 [-] |
| | Total volume (ml), median [IQR] | - | 15 [-] |
| | Missing | - | 6 |
| Opioid added | | | |
| Opioid Added? | Yes | - | 163 / 302 (54%) |
| | Missing | - | 28 |
| Fentanyl | N (%) | - | 112 (37%) |
| Fentanyl Dose (mg/ml) | Median [IQR] | - | 0.100 [0.050 – 0.100] |
| Morphine | N (%) | - | 1 (<1%) |
| Morphine Dose (mg/ml) | Median [IQR] | - | 0.003 [-] |
| Diamorphine | N (%) | - | 50 (17%) |
| Diamorphine Dose (mg/ml) | Median [IQR] | - | 3.0 [2.5 – 3.0] |
| Analgesic Infusion During and After Surgery | | | |
| Local anaesthetic agent | | | |
| Bupivacaine | Used? | - | 62 / 328 (19%) |
| | Concentration (%), median [IQR] | - | 0.125 [0.125 – 0.125] |
| | Total volume (ml), median [IQR] | - | 5 [5 – 6] |
| Levobupivacaine | Used? | - | 264 / 328 (80%) |
| | Concentration (%), median [IQR] | - | 0.125 [0.100 – 0.125] |
| | Total volume (ml), median [IQR] | - | 6 [5 – 8] |
| | Missing | - | 2 |
| Opioid added | | | |
| Opioid Added? | Yes | - | 283 / 317 (89%) |
| | Missing | - | 13 |
| Fentanyl | N (%) | - | 280 / 317 (88%) |
| Fentanyl Dose (mg/ml) | Median [IQR] | - | 2.0 [2.0 – 4.0] |
| | Missing | - | 25 |
| Diamorphine | N (%) | - | 3 / 317 (<1%) |
| Diamorphine | Median [IQR] | - | 3 [-] |

| | | | |
|---|---|---|---|
| Dose (mg/ml) | Missing | - | 2 |
| Further Top Up Required? | N (%) | - | 94 (35%) |
| | Missing | - | 58 |
| Local Analgesia: PVB | | | |
| Were three single shot injections given? | Yes | 316 / 329 (96%) | - |
| | No | 13 / 329 (4%) | - |
| | Missing | 14 | - |
| Technique used | Ultrasound | 141 / 333 (42%) | - |
| | Landmark technique | 192 / 333 (58%) | - |
| | Missing | 10 | - |
| Were catheters subsequently place by surgeon | Yes | 324 / 334 (97%) | - |
| | No | 10 / 334 (3%) | - |
| | Missing | 9 | - |
| Level of insertion of PVB catheter: | Median [IQR] | 5 [4 – 5] | - |
| Opioid Added to maintenance infusion | Yes | 5 / 272 (2%) | - |
| | Missing | 71 | - |
| Level of single-shot Pre-op: median [IQR] | Shot 1 | 4 [4 – 4] | - |
| | Missing | 16 | - |
| | Shot 2 | 6 [5 – 6] | - |
| | Missing | 19 | - |
| | Shot 3 | 7 [6 – 8] | - |
| | Missing | 29 | - |
| Local anaesthetic given: | | | |
| Bupivacaine | Used? | 32 / 271 (12%) | - |
| | Concentration (%), median [IQR] | 0.25 [0.25 – 0.25] | - |
| | Total volume (ml), median [IQR] | 40 [30 – 45] | - |
| Levobupivacaine | Used? | 238 / 271 (88%) | - |
| | Concentration (%), median [IQR] | 0.25 [0.25 - 0.25] | - |
| | Missing | 1 | - |
| | Total volume (ml), median [IQR] | 40 [30 – 45] | - |
| | Missing | 2 | - |
| Ropivacaine | Used? | 1 / 271 (<1%) | - |
| | Concentration (%), median [IQR] | 0.20 [-] | - |
| | Total volume (ml), median [IQR] | 20 [-] | - |
| Opioid added | | | |
| Opioid Added? | Yes | 7 / 273 (3%) | - |
| | Missing | 70 | - |

| | | | |
|---|---|---|---|
| Fentanyl | N (%) | 6 / 273 (2%) | - |
| | Fentanyl Dose (mcg), median [IQR] | 0.09 [0.05 – 0.15] | - |
| Morphine | N (%) | 1 / 273 (<1%) | - |
| | Morphine Dose (mg), median [IQR] | 10 [-] | - |
| Loading Paravertebral Catheters | | | |
| Local anaesthetic given: | | | |
| Bupivacaine | Used? | 30 / 245 (12%) | - |
| | Concentration (%), median [IQR] | 0.25 [0.25-0.25] | - |
| | Total volume (ml), median [IQR] | 20 [15 - 20] | - |
| Levobupivacaine | Used? | 214 / 245 (87%) | - |
| | Concentration (%), median [IQR] | 0.25 [0.25 – 0.25] | - |
| | Missing | 1 | - |
| | Total volume (ml), median [IQR] | 20 [10 – 20] | - |
| | Missing | 2 | - |
| Ropivacaine | Used? | 1 / 245 (<1%) | - |
| | Concentration (%), median [IQR] | 1.0 [-] | - |
| | Total volume (ml), median [IQR] | 5 [-] | - |
| Opioid added | | | |
| Opioid Added? | Yes | 2 / 244 (<1%) | - |
| | Missing | 99 | - |
| Fentanyl | N (%) | 1 / 244 (<1%) | - |
| | Fentanyl Dose (mcg), median [IQR] | 75 [-] | - |
| Morphine | N (%) | 1 / 244 (<1%) | - |
| | Morphine Dose (mg), median [IQR] | 10 [-] | - |
| Analgesia Infusion During and After Surgery | | | |
| Local anaesthetic given: | | | |
| Bupivacaine | Used? | 128 / 281 (46%) | - |
| | Concentration (%), median [IQR] | 0.25 [0.125 – 0.25] | - |
| | Missing | 0 | - |
| | Total volume (ml), median [IQR] | 10 [10 – 15] | - |
| | Missing | 3 | - |
| Levobupivacaine | Used? | 134 / 281 (48%) | - |

| | | | |
|---|---|---|---|
| | Concentration (%), median [IQR] | 0.125 [0.125 – 0.125] | - |
| | Missing | 1 | - |
| | Total volume (ml), median [IQR] | 10 [10 – 15] | - |
| | Missing | 4 | - |
| Ropivacaine | Used? | 8 / 281 (4%) | - |
| | Concentration (%), median [IQR] | 0.20 [0.20 – 0.20] | - |
| Opioid added | | | |
| Opioid Added? | Yes | 5 / 272 (2%) | - |
| | Missing | 71 | - |
| Fentanyl | N (%) | 5 / 272 (2%) | - |
| | Fentanyl Dose (mcg), median [IQR] | 4 [2 – 4] | - |
| | Missing | 2 | - |
| Top-ups Bolus | | | |
| Further Top Ups Required? | N (%) | 28 (10%) | - |
| | Missing | 284 | - |
| **Analgesia on Day of Surgery (Includes pre- and intraoperative analgesia)** | | | |
| Non – Opioid Analgesia | Paracetamol | 320 (88%) | 306 (82%) |
| | NSAIDS (e.g. Diclofenac, Ketorolac) | 106 (29%) | 94 (25%) |
| | Non-opioid Adjunctive Analgesia (e.g gabapentinoids, ketamine) | 48 (13%) | 35 (9%) |
| Opioid Analgesia | Weak Opioids (e.g. Codeine, Tramadol) | 67 (18%) | 78 (21%) |
| | IV Strong Opioid | 324 (89%) | 239 (64%) |
| | Oral Strong Opioid | 47 (13%) | 40 (11%) |
| Oral Morphine Equivalents | Mean (SD)[2] | 83.8 (47.8) | 51.0 (41.3) |
| | Missing | 109 | 164 |
| **Analgesia during Days 1 - 3** | | | |
| Non – Opioid Analgesia | Non-opioid Adjunctive Analgesia (e.g gabapentinoids, ketamine) | 62 (17%) | 53 (14%) |
| Opioid Analgesia | Weak Opioids (e.g. Codeine, Tramadol) | 130 (36%) | 121 (32%) |
| | IV Strong Opioid | 131 (36%) | 41 (11%) |
| | Oral Strong Opioid | 19 (5%) | 21 (6%) |
| Oral Morphine | Mean (SD)[3] | 123.3 (123.2) | 74.2 (84.8) |

| | | | |
|---|---|---|---|
| Equivalents | Missing | 133 | 118 |

[1] Multiple analgesics may have been given.
[2] Adjusted mean difference (adjusted for minimisation parameters) and 95% confidence interval: PVB v TEB = 31.98 (24.14, 39.83) mg. Differences > 0 indicate more analgesia used with PVB.
[3] Adjusted mean difference (adjusted for minimisation parameters) and 95% confidence interval: PVB v TEB = 45.0 (27.6, 62.3) mg. Differences > 0 indicate more analgesia used with PVB.

**Supplementary Table 5: Post-Operation Details**

| | | PVB<br>N=364 | TEB<br>N = 373 |
|---|---|---|---|
| Was the patient discharged within 3 days? | Yes | 67 (19%) | 65 (17%) |
| | No | 294 (81%) | 308 (83%) |
| | Missing | 3 | 0 |
| Post-Operative Ward Care | | | |
| Number of days: General Ward | Median [IQR] | 3 [0 – 5] | 3 [0 – 5] |
| Number of days: Acute Ward | Median [IQR] | 0 [0 – 1] | 0 [0 – 2] |
| Number of days: HDU Level 2 | Median [IQR] | 1 [0 – 3] | 1 [1 – 3] |
| Number of days: ITU Level 3 | Median [IQR] | 0 [0 – 0] | 0 [0 – 0] |
| | Missing | 3 | 0 |
| Total number of days | Median [IQR] | 5 [4 – 9] | 6 [4 – 8] |
| | Missing | 3 | 0 |
| Return to theatre | | | |
| Did the patient return to theatre? | Yes | 21 (6%) | 10 (3%) |
| | No | 329 (94%) | 345 (97%) |
| | Missing | 14 | 10 |
| Reason for return – Day 0[1] | Bronchoscopy | 0 / 5 (-) | 2 / 2 (100%) |
| | Redo thoracotomy | 4 / 5 (80%) | 0 / 2 (-) |
| | Other | 1 / 5 (20%) | 0 / 2 (-) |
| | Missing | 1 | 0 |
| Reason for return – Days 1 – 3[1] | Bronchoscopy | 4 / 9 (44%) | 0 / 3 (-) |
| | Redo thoracotomy | 4 / 9 (44%) | 2 / 3 (67%) |
| | Other | 2 / 9 (22%) | 1 / 3 (33%) |
| | Missing | 0 | 0 |
| Reason for return – Days 4 – Discharge[1] | Bronchoscopy | 4 / 9 (44%) | 3 / 5 (60%) |
| | Redo thoracotomy | 1 / 9 (11%) | 2 / 5 (40%) |
| | Other | 5 / 9 (55%) | 0 / 5 (-) |
| | Missing | 1 | 1 |

[1]Multiple options can be selected

**Supplementary Table 6: Ongoing analgesic management**

| | | PVB<br>N=364 | TEB<br>N = 373 |
|---|---|---|---|
| Management of local anaesthetic block during days 1-3 | | | |
| Additional unplanned involvement of anaesthetist? | Yes | 28 (8%) | 53 (14%) |
| | Missing | 4 | 0 |
| Optimise block? | Yes | 36 (10%) | 50 (13%) |
| | Missing | 3 | 0 |
| Was the PVB/TEB removed within 48hours post-surgery? | Yes | 72 (20%) | 82 (22%) |
| | Missing | 6 | 7 |
| Pain management from post op to discharge | | | |
| Additional unplanned involvement of the pain team? | Yes | 47 (13%) | 58 (16%) |
| | Missing | 2 | 0 |

**Supplementary Table 7: VAS Chronic Pain Responses – Other secondary analyses**

| | PVB | TEB | Estimate (95% CI) | Estimate (95% CI) |
|---|---|---|---|---|
| Worst pain (VAS[1] ≥70) | | | | |
| 3 Month | 20 / 269 (7%) | 18 / 269 (7%) | 0.01 (-0.03, 0.05)[2] | 1.08 (0.59, 1.99)[3] |
| 6 Month | 27 / 272 (10%) | 16 / 292 (5%) | 0.04 (-0.00, 0.08)[2] | 1.74 (0.96, 3.17)[3] |
| 12 Month | 11 / 212 (5%) | 12 / 232 (5%) | -0.00 (-0.03, 0.02)[2] | 0.98 (0.45, 2.13)[3] |
| Worst Pain VAS[1] (median [IQR], n) | | | | |
| 3 Month | 12 [5-34], 269 | 12 [4-32], 269 | 0.5 [-5, 5.5][4] | - |
| 6 Month | 10 [3-31.5], 272 | 8 [3-26], 292 | 2 [-2, 4][4] | - |
| 12 Month | 7 [2-24], 212 | 7 [2-20], 232 | 2 [-2, 3][4] | - |
| Worst Pain (with regards to Chest) Ordered Categories[5] at 3 months | | | | |
| Severe Pain | 20 / 269 (7%) | 18 / 269 (7%) | 1.07 (0.78, 1.47)[6] | - |
| Moderate Pain | 40 / 269 (15%) | 38 / 269 (14%) | | - |
| Mild Pain | 89 / 269 (33%) | 92 / 269 (34%) | | - |
| No Pain | 120 / 269 (45%) | 121 / 269 (45%) | | - |
| Worst Pain (with regards to Chest) Ordered Categories[5] at 6 months | | | | |
| Severe Pain | 27 / 272 (10%) | 16 / 292 (5%) | 1.10 (0.80, 1.50)[6] | - |
| Moderate Pain | 32 / 272 (12%) | 31 / 292 (11%) | | - |
| Mild Pain | 79 / 272 (29%) | 87 / 292 (30%) | | - |
| No Pain | 134 / 272 (49%) | 158 / 292 (54%) | | - |
| Worst Pain (with regards to Chest) Ordered Categories[5] at 12 months | | | | |
| Severe Pain | 11 / 212 (5%) | 12 / 232 (5%) | 0.93 (0.67, 1.31) [6] | - |
| Moderate Pain | 22 / 212 (10%) | 20 / 232 (9%) | | - |
| Mild Pain | 59 / 212 (28%) | 64 / 232 (27%) | | - |
| No Pain | 120 / 212 (57%) | 136 / 232 (59%) | | - |
| Average Pain VAS[1] (median [IQR], n) | | | | |
| 3 Month | 9 [4-26], 269 | 10 [4-22], 269 | 0 [-4, 3][4] | - |
| 6 Month | 7.5 [2-21.5], 272 | 6.5 [3-18], 292 | 1 [-2, 3][4] | - |
| 12 Month | 6 [2-19], 213 | 6 [2-16], 232 | 0 [-2, 2][4] | - |
| Average pain (VAS[1] ≥70) | | | | |
| 3 Month | 9 / 269 (3%) | 7 / 269 (3%) | 0.00 (-0.02, 0.03)[2] | 1.25 (0.47, 3.29)[3] |
| 6 Month | 15 / 272 (6%) | 8 / 292 (3%) | 0.02 (-0.01, 0.05)[2] | 1.95 (0.84, 4.51)[3] |
| 12 Month | 6 / 213 (3%) | 6 / 232 (3%) | 0.01 (-0.01, 0.02)[2] | 1.06 (0.35, 3.17)[3] |
| Average Pain (with regards to Chest) Ordered Categories[5] at 3 months | | | | |
| Severe Pain | 9 / 269 (3%) | 7 / 269 (3%) | 1.17 (0.85, 1.61)[6] | - |
| Moderate Pain | 33 / 269 (12%) | 27 / 269 (10%) | | - |
| Mild Pain | 91 / 269 (34%) | 101 / 269 (38%) | | - |
| No Pain | 136 / 269 (51%) | 134 / 269 (49%) | | - |
| Average Pain (with regards to Chest) Ordered Categories[5] at 6 months | | | | |
| Severe Pain | 15 / 272 (6%) | 8 / 292 (3%) | 1.12 (0.81, 1.55)[6] | - |
| Moderate Pain | 27 / 272 (10%) | 22 / 292 (7%) | | - |
| Mild Pain | 77 / 272 (28%) | 90 / 292 (31%) | | - |

| | | | | |
|---|---|---|---|---|
| No Pain | 153 / 272 (56%) | 172 / 292 (59%) | | - |
| Average Pain (with regards to Chest) Ordered Categories[5] at 12 months | | | | |
| Severe Pain | 6 / 213 (3%) | 6 / 232 (3%) | 0.91 (0.64, 1.30)[6] | - |
| Moderate Pain | 14 / 213 (7%) | 18 / 232 (8%) | | - |
| Mild Pain | 67 / 213 (31%) | 66 / 232 (28%) | | - |
| No Pain | 126 / 213 (59%) | 142 / 232 (61%) | | - |

[1] VAS scored from 0 to 100 with 100 indicating more pain

[2] Estimated Risk Difference adjusted for minimisation variables. Estimated Risk Difference (RD) <0 indicate less pain with PVB.

[3] Estimated Relative Risk adjusted for minimisation variables. For binary outcome, Relative Risk (RR) <1 favours the PVB group.

[4] Unadjusted median difference using bootstrapping methods. Estimated difference < 0 indicated less pain with PVB.

[5] 0-9 ‘no pain’; 10-39 ‘mild pain’; 40-69 ‘moderate pain’; 70-100 ‘severe pain’

[6] Estimated proportional Odd Ratio adjusted for minimisation variables. Proportional Odds Ratio < 0 favours PVB.

**Supplementary Table 8: VAS Chronic Pain Responses – Subgroup Analyses**

| CPTP ≥40 at 6 months | PVB | TEB | Comparison (RR[1]), (95% CI) | P-value for interaction |
|---|---|---|---|---|
| Subgroup: Gender | | | | |
| Male | 31 / 152 (20%) | 27 / 159 (17%) | 1.16 (0.82, 1.63) | 0.40 |
| Female | 28 / 120 (23%) | 20 / 133 (15%) | 1.54 (0.88, 2.69) | |
| Subgroup: Ages (years) | | | | |
| <65 | 28 / 117 (24%) | 25 / 109 (23%) | 1.08 (0.64, 1.81) | 0.23 |
| ≥65 | 31 / 155 (20%) | 22 / 183 (12%) | 1.64 (1.09, 2.47) | |
| Subgroup: Thoracotomy | | | | |
| Lung cancer | 53 / 230 (23%) | 41 / 249 (16%) | 1.37 (0.98, 1.93) | 0.31 |
| Other | 6 / 42 (14%) | 6 / 43 (14%) | 0.92 (0.50, 1.68) | |

[1]RR<1 indicate less pain with PVB

**Supplementary Table 9: VAS Chronic Pain Responses – Sensitivity Analysis**

| | PVB | TEB | Risk Difference[1] (95% CI) | Relative Risk[2] (95% CI) |
|---|---|---|---|---|
| Per Protocol Analysis: CTPT ≥40 at 6 months | | | | |
| Yes | 55 (22%) | 43 (17%) | 0.03 (-0.02, 0.08) | 1.40 (0.85, 2.29) |
| No | 199 (78%) | 208 (83%) | | |
| Missing | 3 | 1 | | |
| Sensitivity Analysis for Sustained Pain: CTPT ≥40 at both 3 months and 6 months | | | | |
| Yes | 33 (12%) | 25 (9%) | 0.04 (-0.02, 0.11) | 1.24 (0.87, 1.77) |
| No | 241 (88%) | 268 (91%) | | |
| Missing | 1 | 0 | | |

**Supplementary Figure 1: Tipping Point Scenario A**

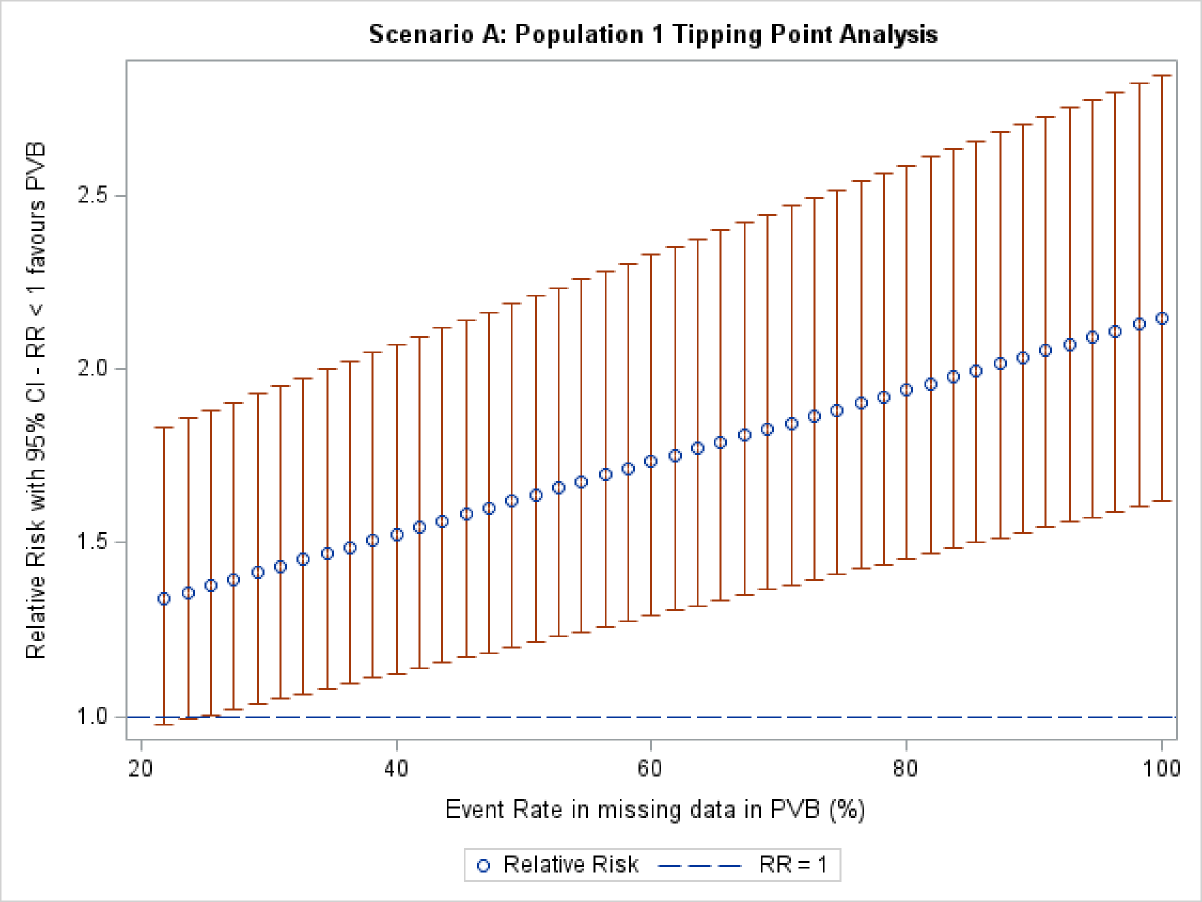

**Supplementary Figure 2: Tipping Point Scenario B**

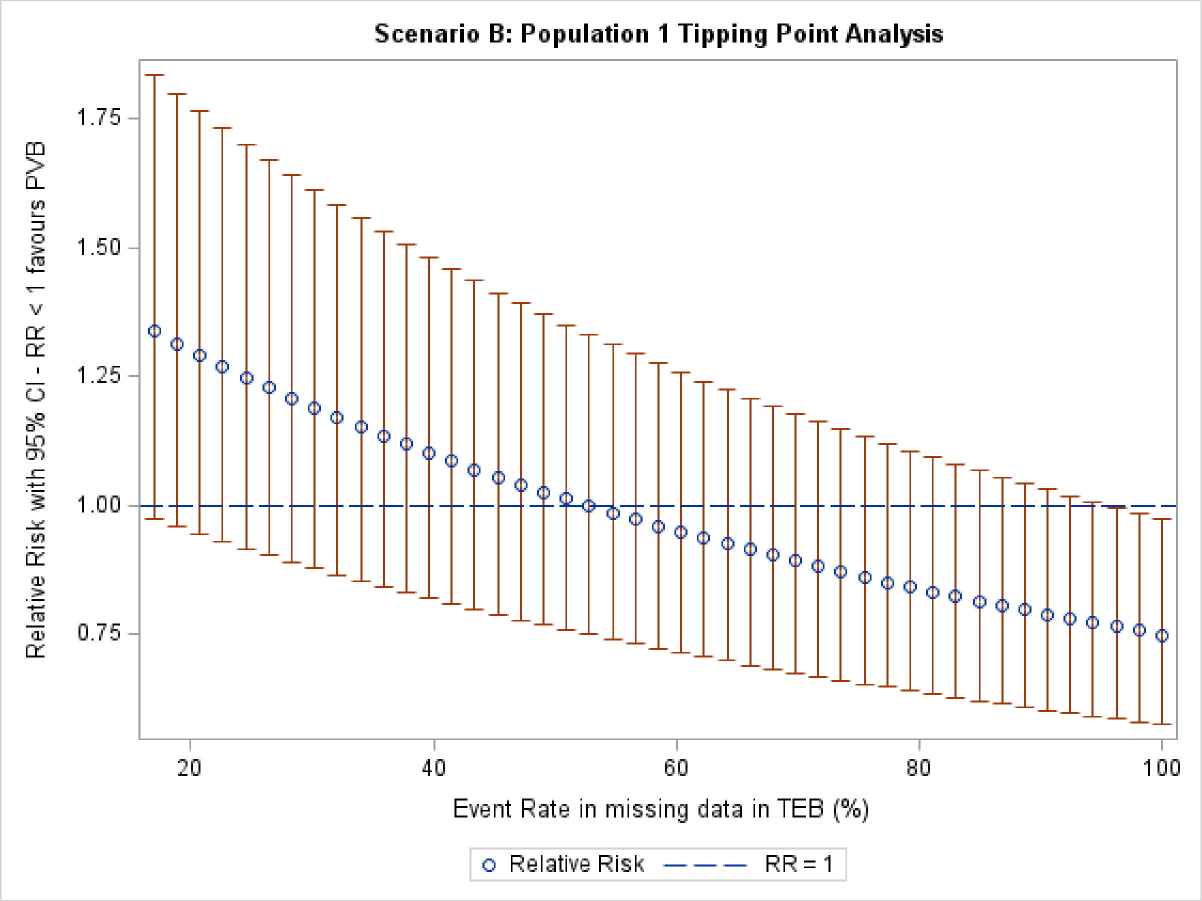

**Supplementary Table 10: Other Chronic Pain and Health Related Quality of Life Responses**

| | PVB | TEB | Adjusted Mean Difference (95% CI) |
|---|---|---|---|
| HADS[1]: Anxiety Subscale (Mean (SD), n) | | | |
| Baseline | 5.61 (4.39), 367 | 5.80 (4.27), 370 | - |
| Discharge | 4.86 (4.20), 252 | 5.32 (4.16), 275 | -0.39 (-0.96, 0.17)[2,3] |
| 3 Month | 5.38 (4.22), 228 | 5.39 (4.35), 220 | 0.40 (-0.23, 1.03) [2,3] |
| 6 Month | 5.33 (4.52), 219 | 5.09 (4.35), 239 | 0.69 (0.03, 1.34) [2,3] |
| 12 Month | 5.45 (4.51), 181 | 5.33 (4.41), 196 | 0.46 (-0.24, 1.16) [2,3] |
| HADS[1]: Depression Subscale (Mean (SD), n) | | | |
| Baseline | 3.51 (3.61), 368 | 3.69 (3.28), 370 | - |
| Discharge | 4.78 (3.82), 252 | 4.85 (3.62), 274 | 0.14 (-0.37, 0.66) [2,3] |
| 3 Month | 5.06 (4.31), 228 | 4.64 (3.60), 220 | 0.67 (0.07, 1.27) [2,3] |
| 6 Month | 4.96 (4.76), 219 | 4.78 (3.98), 239 | 0.64 (-0.04, 1.32) [2,3] |
| 12 Month | 5.01 (4.48), 182 | 4.95 (4.13), 196 | 0.64 (-0.06, 1.35) [2,3] |
| EQ-5D-5L: Total Index Score[6] (Mean (SD), n) | | | |
| Baseline | 0.80 (0.20), 377 | 0.79 (0.19), 374 | - |
| Hospital Discharge | 0.59 (0.22), 255 | 0.61 (0.22), 275 | -0.03 (-0.06, 0.01)[4,5] |
| 3 Month | 0.68 (0.24), 267 | 0.69 (0.23), 269 | -0.01 (-0.04, 0.03) [4,5] |
| 6 Month | 0.67 (0.26), 265 | 0.70 (0.23), 276 | -0.03 (-0.07, 0.00) [4,5] |
| 12 Month | 0.67 (0.28), 213 | 0.73 (0.22), 229 | -0.06 (-0.09, -0.02) [4,5] |
| EQ-5D-5L: Total VAS Score[7] (mean (SD), n) | | | |
| Baseline | 74.79 (19.2), 375 | 73.13 (21.1), 377 | - |
| Hospital Discharge | 65.16 (19.9), 285 | 64.00 (20.2), 308 | 0.56 (-2.44, 3.56) [4,5] |
| 3 Month | 68.56 (21.2), 269 | 70.47 (21.1), 268 | -2.25 (-5.58, 1.08) [4,5] |
| 6 Month | 69.26 (23.8), 269 | 72.53 (19.9), 277 | -3.71 (-7.18, -0.24) [4,5] |
| 12 Month | 71.90 (22.5), 210 | 70.12 (23.8), 229 | 0.89 (-3.12, 4.90) [4,5] |
| SF-MPQ-2[8]: Baseline (mean (SD), n) | | | |
| Total Pain Score | 1.39 (3.56), 342 | 1.22 (3.09), 355 | - |
| Continuous pain | 0.44 (1.14), 342 | 0.38 (0.91), 355 | - |
| Intermittent pain | 0.32 (1.04), 343 | 0.29 (0.93), 355 | - |
| Predominantly neuropathic pain | 0.24 (0.80), 343 | 0.20 (0.73), 355 | - |
| Affective pain | 0.45 (1.32), 343 | 0.35 (0.99), 355 | - |
| SF-MPQ-2[8]: 3 months (mean (SD), n) | | | |
| Total Pain Score | 4.53 (6.33), 232 | 3.88 (5.38), 233 | 0.57 (-0.46, 1.60)[9] |
| Continuous pain | 1.30 (1.80), 232 | 1.04 (1.57), 235 | 0.27 (-0.03, 0.57) [9] |
| Intermittent pain | 1.06 (1.82), 232 | 0.97 (1.64), 237 | 0.07 (-0.23, 0.38) [9] |
| Predominantly neuropathic pain | 1.05 (1.55), 232 | 1.01 (1.44), 236 | 0.03 (-0.23, 0.29) [9] |

| | | | |
|---|---|---|---|
| Affective pain | 1.20 (1.98), 235 | 1.02 (1.76), 236 | 0.18 (-0.15, 0.51) [9] |
| SF-MPQ-2[8]: 6 months (mean (SD), n) | | | |
| Total Pain Score | 3.70 (6.09), 240 | 2.92 (4.60), 237 | 0.77 (-0.14, 1.68) [9] |
| Continuous pain | 1.09 (1.73), 241 | 0.74 (1.27), 237 | 0.36 (0.10, 0.62) [9] |
| Intermittent pain | 0.83 (1.67), 240 | 0.75 (1.54), 238 | 0.06 (-0.22, 0.33) [9] |
| Predominantly neuropathic pain | 0.88 (1.50), 241 | 0.78 (1.21), 238 | 0.13 (-0.10, 0.36) [9] |
| Affective pain | 1.00 (1.97), 242 | 0.73 (1.32), 239 | 0.23 (-0.06, 0.52) [9] |
| SF-MPQ-2[8]: 12 months (mean (SD), n) | | | |
| Total Pain Score | 3.83 (6.61), 182 | 2.81 (4.78), 196 | 0.85 (-0.22, 1.93) [9] |
| Continuous pain | 1.23 (2.02), 184 | 0.83 (1.51), 198 | 0.38 (0.05, 0.71) [9] |
| Intermittent pain | 0.78 (1.76), 184 | 0.67 (1.43), 199 | 0.08 (-0.22, 0.39) [9] |
| Predominantly neuropathic pain | 0.91 (1.54), 182 | 0.73 (1.28), 197 | 0.10 (-0.17, 0.36) [9] |
| Affective pain | 1.01 (1.87), 184 | 0.68 (1.36), 201 | 0.33 (0.03, 0.63) [9] |
| BPI[10]: Pain Interference Score (mean (SD), n) | | | |
| Baseline | 1.26 (2.05), 322 | 1.37 (2.14), 340 | - |
| 3 Month | 2.61 (2.58), 268 | 2.28 (2.33), 269 | 0.24 (-0.16, 0.65) [9] |
| 6 Month | 2.48 (2.76), 269 | 2.14 (2.39), 278 | 0.33 (-0.09, 0.75) [9] |
| 12 Month | 2.41 (2.77), 211 | 1.95 (2.19), 230 | 0.40 (-0.03, 0.83) [9] |
| Patient Satisfaction | | | |
| Discharge | PVB (N=354) | TEB (N=363) | Adjusted Proportional Odds Ratio[11] (95% CI) |
| How satisfied have you been with pain therapy after surgery? (N(%)) | | | |
| Very satisfied | 159 (55%) | 177 (57%) | 0.83 (0.60, 1.14) |
| Satisfied | 116 (40%) | 112 (36%) | |
| Dissatisfied | 11 (4%) | 8 (3%) | |
| Very dissatisfied | 3 (1%) | 11 (4%) | |
| Missing | 65 | 55 | |
| How satisfied were you with the care provided by the hospital in general? (N(%)) | | | |
| Very satisfied | 231 (80%) | 253 (82%) | 0.85 (0.56, 1.27) |
| Satisfied | 54 (19%) | 44 (14%) | |
| Dissatisfied | 1 (<1%) | 6 (2%) | |
| Very dissatisfied | 3 (1%) | 5 (2%) | |
| Missing | 65 | 55 | |
| 3 Month | PVB (N=269) | TEB (N=271) | Adjusted Proportional Odds Ratio[11] (95% CI) |
| How satisfied have you been with pain therapy after surgery? (N(%)) | | | |
| Very satisfied | 136 (51%) | 145 (54%) | 0.87 (0.63, 1.21) |
| Satisfied | 108 (41%) | 106 (39%) | |
| Dissatisfied | 17 (6%) | 11 (4%) | |
| Very dissatisfied | 4 (2%) | 7 (3%) | |

| | | | |
|---|---|---|---|
| Missing | 5 | 2 | |
| How satisfied were you with the care provided by the hospital in general? (N(%)) | | | |
| Very satisfied | 183 (69%) | 198 (74%) | 0.79 (0.55, 1.13) |
| Satisfied | 70 (26%) | 59 (22%) | |
| Dissatisfied | 8 (3%) | 6 (2%) | |
| Very dissatisfied | 6 (2%) | 5 (2%) | |
| Missing | 3 | 3 | |
| 6 Months | N = 276 | N = 294 | Adjusted Proportional Odds Ratio[11] (95% CI) |
| How satisfied have you been with pain therapy after surgery? (N(%)) | | | |
| Very satisfied | 141 (53%) | 166 (59%) | 0.78 (0.56, 1.08) |
| Satisfied | 109 (41%) | 101 (36%) | |
| Dissatisfied | 13 (5%) | 10 (4%) | |
| Very dissatisfied | 3 (1%) | 4 (1%) | |
| Missing | 9 | 12 | |
| How satisfied were you with the care provided by the hospital in general? (N(%)) | | | |
| Very satisfied | 190 (71%) | 213 (76%) | 0.87 (0.60, 1.25) |
| Satisfied | 66 (25%) | 57 (20%) | |
| Dissatisfied | 5 (2%) | 8 (3%) | |
| Very dissatisfied | 6 (2%) | 4 (1%) | |
| Missing | 8 | 11 | |
| 12 Month | N = 221 | N = 237 | Adjusted Proportional Odds Ratio[11] (95% CI) |
| How satisfied have you been with pain therapy after surgery? (N(%)) | | | |
| Very satisfied | 116 (55%) | 149 (64%) | 0.64 (0.45, 0.92) |
| Satisfied | 82 (39%) | 72 (31%) | |
| Dissatisfied | 9 (4%) | 4 (2%) | |
| Very dissatisfied | 3 (1%) | 7 (3%) | |
| Missing | 11 | 5 | |
| How satisfied were you with the care provided by the hospital in general? (N(%)) | | | |
| Very satisfied | 161 (76%) | 178 (76%) | 0.96 (0.64, 1.42) |
| Satisfied | 41 (20%) | 48 (21%) | |
| Dissatisfied | 5 (2%) | 3 (1%) | |
| Very dissatisfied | 4 (2%) | 4 (2%) | |
| Missing | 10 | 4 | |

[1] HADs subscales scored from 0 to 21 with higher scores implying worse state.

[2] For continuous Scores mean difference <0 indicate less anxiety/depression with PVB

[3] Adjusted for minimisation variables including Baseline score. Estimated difference <0 indicate less anxiety/depression with PVB.

[4] For EQ5D Index and VAS score mean difference >0 indicate better overall health with PVB

[5] Adjusted for minimisation variables including Baseline score. Time and Time*Group interaction. Estimated difference between groups >0 indicate better overall health with PVB.

[6] EQ-5D-5L index score from -0.594 to 1 with 1 implying perfect health and negatives scores a state worse than death
[7] EQ-5D-5L VAS scored from 0 – 100 with higher scores implying better health.
[8] SF-MPQ-2 domains scored from 0 – 11 with higher numbers indicating worse pain. Total pain scored 0-44.
[9] Adjusted for minimisation variables, Time and Time*Group interaction. Estimated difference between groups <0 indicate less pain with PVB.
[10] BPI scored from 0 – 10 with 10 indicating worse pain.
[11] Adjusted for minimisation variables. Proportional odds ratios > 1 prefers PVB.

**Supplementary Table 11: Acute Pain Responses**

| | PVB<br>Mean (SD), n | TEB<br>Mean (SD), n | Adjusted Mean Difference[2,3] (95% CI) |
|---|---|---|---|
| Worst Pain (with regards to chest) VAS (mm) | | | |
| Baseline | 8.9 (18.5), 362 | 9.1 (19.4), 369 | - |
| Day 1 | 64.8 (29.2), 318 | 57.6 (34.4), 331 | 7.7 (2.8, 12.5) |
| Day 2 | 61.6 (29.4), 296 | 57.9 (29.3), 307 | 3.6 (-1.0, 8.2) |
| Day 3 | 56.9 (30.2), 255 | 54.2 (31.0), 274 | 1.5 (-3.5, 6.5) |
| Discharge | 48.1 (30.0), 291 | 46.9 (28.6), 309 | 1.3 (-3.3, 5.9) |
| Average Pain (with regards to chest) VAS (mm) | | | |
| Baseline | 6.1 (13.4), 362 | 6.2 (14.4), 369 | - |
| Day 1 | 46.1 (26.9), 318 | 39.1 (29.5), 332 | 7.0 (2.7, 11.2) |
| Day 2 | 43.7 (26.3), 296 | 40.1 (26.3), 307 | 3.3 (-0.8, 7.4) |
| Day 3 | 40.5 (27.4), 255 | 37.0 (26.8), 275 | 2.8 (-1.6, 7.3) |
| Discharge | 31.1 (22.8), 291 | 29.4 (22.1), 308 | 1.9 (-1.6, 5.4) |
| Total Pain over Acute Phase VAS Score[1] | | | |
| Days 1-3 | 129.3 (62.3), 229 | 115.6 (59.6), 247 | 13.5 (2.9, 24.0) |
| BPI – Pain Interference Score[4] | | | |
| Baseline | 1.26 (2.05), 322 | 1.37 (2.14), 340 | - |
| Day 1 | 4.57 (2.76), 318 | 3.97 (2.90), 324 | 0.63 (0.21, 1.05) |
| Day 2 | 4.50 (2.63), 292 | 4.13 (2.74), 300 | 0.32 (-0.09, 0.74) |
| Day 3 | 4.31 (2.73), 255 | 3.92 (2.78), 270 | 0.35 (-0.09, 0.78) |
| Hospital Discharge | 3.32 (2.42), 288 | 3.16 (2.33), 309 | 0.15 (-0.21, 0.51) |

| | | | |
|---|---|---|---|
| SF-MPQ-2[5] at Baseline | | | |
| Total Pain Score | 1.39 (3.56), 342 | 1.22 (3.09), 355 | - |
| Continuous pain | 0.44 (1.14), 342 | 0.38 (0.91), 355 | - |
| Intermittent pain | 0.32 (1.04), 343 | 0.29 (0.93), 355 | - |
| Predominantly neuropathic pain | 0.24 (0.80), 343 | 0.20 (0.73), 355 | - |
| Affective pain | 0.45 (1.32), 343 | 0.35 (0.99), 355 | - |
| SF-MPQ-2[5] at Discharge | | | |
| Total Pain Score | 7.60 (6.79), 269 | 7.09 (6.90), 284 | 0.55 (-0.57, 1.67) |
| Continuous pain | 2.49 (2.07), 270 | 2.41 (2.13), 291 | 0.09 (-0.24, 0.43) |
| Intermittent pain | 2.11 (2.23), 269 | 1.96 (2.13), 286 | 0.17 (-0.19, 0.53) |
| Predominantly neuropathic pain | 1.07 (1.50), 269 | 1.15 (1.60), 286 | -0.08 (-0.33, 0.18) |
| Affective pain | 1.97 (2.21), 271 | 1.80 (2.27), 290 | 0.20 (-0.17, 0.56) |

[1] VAS scored from 0 – 100 with 0 indicating no pain and 100 indicating severe pain.

[2] Group Difference <0 indicate less pain with PVB.

[3] Adjusted for minimisation variables, Time and Time*Group interaction: Group Difference <0 favours the PVB group

[4] BPI scored from 0 to 10 with higher scores indicating more pain

[5] SF-MPQ-2 domains are scored from 0 to 11 with higher scores indicating worse state. Total pain is scored from 0 – 44.

**Supplementary Table 12: Long-term analgesic use**

| | PVB N / N (%) | TEB N / N (%) | Adjusted Risk Difference[1](95% CI) | Adjusted Relative Risk[2] (95% CI) |
|---|---|---|---|---|
| Sustained Long Term Medication Use (at least once a week) | | | | |
| Conventional Pain Killers | | | | |
| 3 Month | 186 / 310 (60%) | 173 / 315 (55%) | 0.05 (-0.03, 0.13) | 1.10 (0.96, 1.26) |
| 6 Month | 41 / 297 (14%) | 43 / 313 (14%) | -0.00 (-0.06, 0.05) | 0.99 (0.67, 1.48) |
| 12 Month | 34 / 256 (13%) | 31 / 275 (11%) | 0.00 (-0.05, 0.06) | 1.18 (0.75, 1.85) |
| Opioids | | | | |
| 3 Month | 155 / 306 (51%) | 158 / 317 (50%) | 0.01 (-0.07, 0.08) | 1.03 (0.89, 1.21) |
| 6 Month | 31 / 291 (11%) | 34 / 313 (11%) | -0.01 (-0.05, 0.03) | 0.97 (0.61, 1.52) |
| 12 Month | 20 / 253 (8%) | 25 / 274 (9%) | -0.03 (-0.07, 0.01) | 0.86 (0.49, 1.49) |
| Neuropathic Painkillers | | | | |
| 3 Month | 48 / 291 (16%) | 41 / 302 (14%) | 0.03 (-0.02, 0.09) | 1.18 (0.81, 1.73) |
| 6 Month | 20 / 291 (7%) | 27 / 311 (9%) | -0.01 (-0.04, 0.02) | 0.75 (0.44, 1.30) |
| 12 Month | 20 / 253 (8%) | 20 / 272 (7%) | 0.01 (-0.03, 0.05) | 1.01 (0.56, 1.83) |
| Anti-inflammatory Drugs | | | | |
| 3 Month | 30 / 291 (10%) | 17 / 296 (6%) | 0.58 (0.01, 1.14) | 1.79 (1.01, 3.14) |
| 6 Month | 7 / 290 (2%) | 3 / 309 (<1%) | N/A | N/A |
| 12 Month | 7 / 250 (3%) | 1 / 270 (<1%) | N/A | N/A |
| Long Term Medication Use - Any | | | | |
| Conventional Pain Killers | | | | |
| 3 Month | 176 / 310 (57%) | 163 / 315 (52%) | 0.05 (-0.03, 0.13) | 1.10 (0.95, 1.27) |
| 6 Month | 40 / 297 (13%) | 40 / 313 (13%) | 0.00 (-0.05, 0.06) | 1.04 (0.69, 1.56) |
| 12 Month | 34 / 256 (13%) | 30 / 275 (11%) | 0.01 (-0.04, 0.06) | 1.22 (0.77, 1.92) |
| Opioids | | | | |
| 3 Month | 148 / 306 (48%) | 148 / 317 (47%) | 0.01 (-0.06, 0.09) | 1.06 (0.90, 1.24) |
| 6 Month | 31 / 291 (11%) | 31 / 313 (10%) | -0.00 (-0.05, 0.04) | 1.06 (0.66, 1.69) |

| | | | | |
|---|---|---|---|---|
| 12 Month | 20 / 253 (8%) | 24 / 274 (9%) | -0.03 (-0.07, 0.02) | 0.89 (0.51, 1.56) |
| Neuropathic Painkillers | | | | |
| 3 Month | 48 / 291 (16%) | 36 / 302 (12%) | 0.05 (-0.00, 0.10) | 1.34 (0.90, 1.99) |
| 6 Month | 19 / 291 (7%) | 27 / 311 (9%) | -0.01 (-0.05, 0.02) | 0.72 (0.41, 1.25) |
| 12 Month | 20 / 253 (8%) | 19 / 272 (7%) | 0.01 (-0.03, 0.05) | 1.06 (0.58, 1.94) |
| Anti-inflammatory Drugs | | | | |
| 3 Month | 29 / 291 (10%) | 15 / 296 (5%) | 0.04 (0.01, 0.07) | 1.93 (1.07, 3.50) |
| 6 Month | 6 / 290 (2%) | 3 / 309 (<1%) | N/A | N/A |
| 12 Month | 5 / 250 (2%) | 1 / 270 (<1%) | N/A | N/A |

[1] Adjusted for minimisation variables. Estimated Risk Difference (RD) <0 indicate less medication use with PVB.

[2] Adjusted for minimisation variables. For binary outcome, Relative Risk (RR) <1 favours the PVB group.

### Other Acknowledgements

The authors wish to thank:

- Independent members of the (1) Trial Steering Group and (2) Data Monitoring Committee who oversaw this study:

  1. Anne Devrell (Patient and Public representative), Christine Roffe (Chair, Stroke Physician, Keele University), Michael Shackcloth (Thoracic Surgeon, Liverpool Heart and Chest Hospital NHS Foundation Trust), Laura Bojke (Health Economist, University of York), Andrew Klein (Macintosh Professor of Anaesthesia, Royal Papworth Hospital NHS Trust)

  2. Nadine Foster (Physiotherapist, Keele University), Paul Myles (Clinician, Alfred Hospital, Melbourne) David Cooper (Statistician, University of Aberdeen)

- Participants: The authors also gratefully acknowledge the support of the patients who consented and were randomised into the TOPIC-2 trial, as well as those who were approached and considered participation in TOPIC-2.

### TOPIC-2 Investigators

- Members of the Trial Management Group:

Fang Gao Smith (Chief Investigator), Afreen Khan (Trial Manager), Lee Middleton (Lead Statistician), Hannah Summers (Statistician), Erum Khan (Data Manager), Amy Kerr (Senior Thoracic Surgery Research Nurse), Babu Naidu (Associate Professor of Cardiothoracic Surgery) Andreas Goebel (Senior Lecturer in Pain Medicine), Teresa Melody (Chair of Clinical Research Ambassador Group), Joyce Yeung (West Midlands Trainee Research in Anaesthesia and Intensive Care Network (WMTRAIN) representative), Sarah Tearne (Clinical Trials Team Leader), Louise Jackson (Lecturer in Health Economics), Matt Wilson (Clinical Scientist in Anaesthesia), Marcus Jepson (Lecturer in Qualitative Health Science), Nandor Marczin (Honorary Consultant Anaesthetist), Ben Shelley (Consultant in Cardiothoracic Anaesthesia), Lajos Szentgyorgyi (Consultant Anaesthetist), Andrew Worrall (Patient representative), Stephen Grant (Patient representative ).

- Other Current and Former BCTU Staff

Ben Watkins (Senior Trial Manager), Jordan Evans (Data Manager), Hugh Jarret (Trial Management Team Lead), Rajnikant Mehta (Statistician), BCTU Programming Team

- TOPIC-2 Recruiting sites:

| **Site** | **Staff** |
|---|---|
| **Castle Hill Hospital** | Syed Qadri (PI), Karen Dobbs |
| **University Hospital Coventry** | Mathew Varghese Patteril (PI), Dawn Davies |
| **Essex Cardiothoracic Centre** | Gyanesh J. Namjoshi (PI), Sofia Matias, Jonaifah Ramirez |
| **Glenfield Hospital** | Sridhar Rathinam (PI), Rebecca Boyles |
| **Golden Jubilee Hospital** | Ben Shelley (PI), Philip McCall, Mark Thornton, Charlene Hamilton, Elizabeth Boyd, Christine Aitken, Julie Buckley |
| **Harefield Hospital** | Nandi Marczin (PI), Marita Quiroz-Patel |
| **Heartlands Hospital**<br>**Queen Elizabeth Hospital** | Babu Naidu (PI), Chris Lacson, Aya Osman |
| **Nottingham City Hospital** | Munib Malak (PI), Adele Malson, Joanna Curtis |

| | |
|---|---|
| **Wythenshawe Hospital** | Lajos Szentgyorgyi (PI), Poppy Froggett, Pamela Maroa, Clinton Prakash |
| **Morriston Hospital** | Michael Gilbert (PI), Jenny Travers |
| **Belfast Royal Victoria** | David Johnston (PI), Christine Turley |
| **Aberdeen Royal Infirmary** | John Chalmers (PI), Sandra Mann |
| **Blackpool Royal Victoria** | Petr Martinovsky (PI), Deepa Sebastian, Louie Garcia |
| **St. Barts Hospital** | Sibtain Anwar (PI), Michelle Lee |
| **John Radcliffe Hospital** | Elizabath Belcher (PI), Penny Carter |

UNIVERSITY OF BIRMINGHAM | BCTU Birmingham Clinical Trials Unit

**A Randomised Controlled Trial to investigate the effectiveness of ThOracic Epidural and Paravertebral Blockade In reducing Chronic Post- Thoracotomy Pain: 2**

**The TOPIC 2 Trial**

TOPIC 2

Trial Registration: ISRCTN 45041624

**Statistical Analysis Plan**

| SAP Version Number | Protocol Version Number |
|---|---|
| 1.0 | 6.0a |

| | | | | | |
|---|---|---|---|---|---|
| Name of Author: | Hannah Summers | Role: | Trial Statistician | Affiliation: | BCTU |
| Signature of Author: | H. Summers (email) | Date: | 04/07/2024 | | University of Birmingham |
| Name of Reviewer: | Versha Cheed | Role: | Senior Statistician | Affiliation: | BCTU |
| Signature of Reviewer: | | Date: | 04/07/2024 | | University of Birmingham |
| Name of Chief Investigator: | Prof Fang Gao Smith | Role: | Chief Investigator | Affiliation: | University of Birmingham |
| Signature of Chief Investigator: | | Date: | 04.07.2024 | | |
| **This Statistical Analysis Plan has been approved by:** | | | | | |
| Name of Approver: | Lee Middleton | Role: | Senior Statistician | Affiliation: | BCTU |

| | | | | | |
|---|---|---|---|---|---|
| Signature of Approver: | Lee Middleton | Date: | 10/7/2024 | | University of Birmingham |

Abbreviations & Definitions

| Abbreviation / Acronym | Meaning |
|---|---|
| ACTA | Association of Cardiothoracic Anaesthetists |
| AE | Adverse Event |
| ARDS | Acute Respiratory Distress Syndrome |
| BCTU | Birmingham Clinical Trials Unit |
| BPI | Brief Pain Inventory |
| CI | Confidence Intervals |
| CONSORT | Consolidated Standards of Reporting Trials |
| CPTP | Chronic Post-Thoracotomy Pain |
| CRAG | Clinical Research Ambassador Group |
| CRF | Case Report Form |
| DMC | Data Monitoring Committee |
| EQ-5D-5L | Euroqol questionnaire |
| ESTS | European Society of Thoracic Surgeons |
| GCP | Good Clinical Practice |
| GEE | Generalised Estimating Equations |
| GP | General Practitioner |
| HADS | Hospital Anxiety and Depression Scale |
| HDU | High Dependency Unit |
| HEFT | Heart of England NHS Foundation Trust |
| HRA | Health Research Authority |
| ICF | Informed Consent Form |
| ICH | International Conference on Harmonisation |
| ICU | Intensive Care Unit |
| ISRCTN | International Standard Randomised Controlled Trial Number |
| ITT | Intention to Treat |

MITT Modified Intention to Treat

NHS National Health Service

PCA Patient Controlled Analgesia

PI Principal Investigator

PIS Participant Information Sheet

PoP Post-operative Pneumonia

PPCs Post-operative Pulmonary Complications

PPI Patient and Public Involvement

PSC Post-operative Surgical Complications

PVB Paravertebral Blockade

QA Quality Assurance

QRI Quintet Recruitment Intervention

RCT Randomised Controlled Trial

REC Research Ethics Committee

R&D Research and Development

SAE Serious Adverse Event

SAP Statistical Analysis Plan

SAR Serious Adverse Reaction

SF-MPQ-2 Short Form McGill Pain Score questionnaire

SUSAR Suspected Unexpected Serious Adverse Reaction

TEB Thoracic Epidural Blockade

TMG Trial Management Group

TMM Thoracic Morbidity and Mortality

TSC Trial Steering Committee

UK United Kingdom

UoB University of Birmingham

VAS Visual Analogue Scale

TABLE OF CONTENTS

1. Introduction 8

2. Background and rationale 8

3. Trial objectives 8

4. Trial methods 9

4.1. Trial design 9

4.2. Trial interventions 9

4.3. Primary outcome measure 9

4.4. Secondary outcome measures 10

4.5. Timing of outcome assessments 11

4.6. Randomisation 11

4.7. Sample size 12

4.8. Framework 12

4.9. Interim analyses and stopping guidance 12

4.10. Internal Pilot Progression Rules 13

4.11. Timing of final analysis 13

4.12. Timing of other analyses 14

4.13. Trial comparisons 14

5. Statistical Principles 14

5.1. Confidence intervals and p-values 14

5.2. Adjustments for multiplicity 14

5.3. Analysis populations 14

5.4. Definition of adherence 15

5.5. Handling protocol deviations 15

5.6. Unblinding 15

6. Trial population 16

6.1. Recruitment 16

6.2. Baseline characteristics 16

7. Intervention(s) 16

7.1. Description of the intervention(s) 16
7.2. Adherence to allocated intervention 16
8. Protocol deviations 16
9. Analysis methods 16
9.1. Covariate adjustment 17
9.2. Distributional assumptions and outlying responses 17
9.3. Handling missing data 17
9.4. Analysis methods – primary outcome(s) 17
9.5. Analysis methods – secondary outcomes 18
9.6. Analysis methods – exploratory outcomes and analyses 19
9.7. Safety data 19
9.8. Planned subgroup analyses 20
9.9. Sensitivity and supportive analyses 20
10. Analysis of sub-randomisations 22
11. Health economic analysis 22
12. Statistical software 22
13. References 22

### 1. Introduction

This document is the Statistical Analysis Plan (SAP) for the TOPIC 2 trial and should be read in conjunction with the current trial protocol. This SAP details the proposed analyses and presentation of the data for the main paper(s) reporting the results for the TOPIC 2 trial.

The results reported in these papers will follow the strategy set out here. Subsequent analyses of a more exploratory nature will not be bound by this strategy, though they are expected to follow the broad principles laid down here. The principles are not intended to curtail exploratory analysis (e.g. to decide cut-points for categorisation of continuous variables), nor to prohibit accepted practices (e.g. transformation of data prior to analysis), but they are intended to establish rules that will be followed, as closely as possible, when analysing and reporting data.

Any deviations from this SAP will be described and justified in the final report or publication of the trial (using a table as shown in Appendix A). The analysis will be carried out by an appropriately qualified statistician, who should ensure integrity of the data during their data cleaning processes.

### 2. Background and rationale

In brief, TOPIC 2 is a Multicentre Randomised Controlled Trial investigating the effectiveness of Thoracic Epidural Blockade (TEB) and Paravertebral Blockade (PVB) in reducing Chronic Post-Thoracotomy Pain (CPTP). See the TOPIC2 protocol for further details on the background and rationale. [1-6]

### 3. Trial objectives

The primary objective is to test the hypothesis that in adult patients undergoing elective open thoracotomy, the use of PVB for peri-operative pain relief reduces the presence of chronic pain at six months by at least 10% compared with TEB.

Secondary objectives are as follows:

- To compare the effectiveness of PVB versus TEB in terms of quality of life, neuropathic pain symptoms, symptoms of anxiety/depression and patients' satisfaction up to 12 months following surgery.
- To compare the effectiveness of PVB versus TEB in terms of acute pain control up to 72 hours following surgery, incidence of post-operative major and minor complications and length of post-operative hospital stay.

- To analyse the costs and effectiveness of PVB compared with TEB.

The plan for the cost effectiveness analysis will be detailed elsewhere.

### 4. Trial methods

#### 4.1. Trial design

TOPIC 2 is a prospective multi-centre randomised, open label, parallel group, superiority trial of 770 adult (≥18 years old) thoracotomy patients. Patients will be randomised in a 1:1 equal ratio to receive TEB (standard treatment) or PVB (interventional treatment). See Appendix B for trial schema.

Participants will be recruited from secondary or tertiary care.

Due to the nature of the intervention, it is not possible to blind participants or surgeons.

This trial has an internal pilot phase (see section 4.10).

#### 4.2. Trial interventions

Two existing peri-operative analgesic techniques:

i) PVB: three pre-incisional injections followed by a catheter placement.

ii) TEB placed pre-incision: usual practice.

#### 4.3. Primary outcome measure

The primary outcome is presence of CPTP at 6 months post-randomisation. Participants will be asked to indicate their 'worst chest pain over the last week' on a visual analogue scale (VAS; 0-100). A CPTP event will be taken to be a score greater or equal to 40 indicating at least a moderate level of pain. See Appendix D on data manipulations for how the primary outcome will be derived.

#### 4.4. Secondary outcome measures

The secondary outcomes are as follows (measured at 3, 6, 12 months post randomisation. For more detail, please refer to TOPIC 2 protocol section 8.2 and Table 1 below):

- Complications of regional analgesia (failure of blockade, hypotension (systolic blood pressure (<90mmHg)), inadequate pain relief, low respiratory rate (<10/minute), drowsiness, nausea and vomiting, urinary retention, itching, high block, post-dural puncture headache, vascular puncture, pleural puncture).
- Occurrence and severity of surgical complications until discharge from hospital (occurrence as defined by the European Society of Thoracic Surgeons dataset and severity as defined by the Thoracic Morbidity and Mortality (TMM) classification
- Post-operative pulmonary complications (PPCs) until discharge from hospital
- Critical care admission (levels 2 and 3)
- Mortality (reported for all deaths due to all causes)
- Analgesic use
- Acute pain in the 3 days following surgery and at discharge, completed by the patient (via VAS; Brief Pain Inventory (BPI))
- Pain at discharge from hospital (via VAS, BPI and Short Form McGill questionnaire (SF-MPQ-2))
- Chronic pain (via VAS, BPI and SF-MPQ-2, completed by the participant at 3, 6 and 12 months post randomisation)
- Presence of severe pain at its worst (VAS>=70) in chronic phase at 3, 6 and 12 months
- Presence of moderate pain at its worst (VAS>=40) in chronic phase at 3 and 12 months
- Presence of severe pain on average (VAS>=70) in chronic phase at 3, 6 and 12 months
- Presence of moderate pain on average (VAS>=40) in chronic phase at 3, 6 and 12 months
- General health-related quality of life (by EQ-5D-5L, completed by the participant at discharge and at 3, 6 and 12 months)
- Mental health state (measured by Hospital Anxiety and Depression Scale (HADS), completed by the participant at discharge and at 3, 6 and 12 months).
- Patient satisfaction (by Likert scale, completed by the participant at discharge and at 3, 6 and 12 months)
- Serious Adverse Events (SAEs)

### Table 1 Data collection tools and corresponding outcomes:

| Collection tool | Outcome | Possible responses |
|---|---|---|
| Pain VAS [7] | Chronic phase:<br>• Worst chest pain over the last week score<br>• Average chest pain over the last week score<br>Acute phase:<br>• Worst chest pain over the last 24 hours score<br>• Average chest pain over the last 24 hours score | All 0-100 (higher=worse score) |
| BPI [8] | Interference score | 0-10 (higher=worse score) |
| SF-MPQ-2 [9] | • Continuous pain subscale score<br>• Intermittent pain subscale score<br>• Neuropathic pain subscale score<br>• Affective pain subscale score<br>• Overall score | All 0-10 (higher=worse score) |
| Generic health related quality of life questionnaire (EQ-5D-5L) [10] | • Index score<br>• Thermometer score | • (-0.59=worst outcome, 1.0=best outcome)<br>• (0-100, higher=better) |
| HADS [11] | • Depression score<br>• Anxiety score | Both 0-21 (lower=better) |
| Likert Scale to assess satisfaction | • Satisfaction with pain therapy after surgery<br>• Satisfaction with care provided by hospital | Very dissatisfied/ Dissatisfied/ Satisfied/ Very satisfied |

See Appendix D on data manipulations for how secondary outcomes will be derived.

#### 4.5. Timing of outcome assessments

The schedule of trial procedures and outcome assessments are given in Appendix C.

#### 4.6. Randomisation

Participants will be randomised in a 1:1 ratio to either TEB or PVB.

Randomisation will be provided by a secure online randomisation system at the Birmingham Clinical Trials Unit (BCTU) using a minimisation algorithm will be used within the online randomisation system to ensure balance in the treatment allocation over the following variables:

- Gender
- Age <65 years or ≥65 years
- Centre
- Thoracotomy for lung cancer resection or for other indication

A 'random element' will be included in the minimisation algorithm, so that each participant has a probability (unspecified here), of being randomised to the opposite intervention that they would have otherwise received. Full details of the algorithm used will be stored in a confidential document at BCTU.

### 4.7. Sample size

Assuming a 30% incidence of CPTP in the TEB group (similar to that seen in our previous TOPIC-pilot results [12] and systematic review [13], 294 patients in each group will give 80% power (two-sided p=0.05) to detect a 10% absolute reduction (i.e. down to 20%, a 33% relative reduction) in the PVB group. Assuming a 10% rate of death (similar to that seen in TOPIC) and a further 15% loss to follow-up at 6 months we will recruit 770 participants.

Our survey of practise of consultant thoracic anaesthetists at UK thoracic centres indicated that a 50% relative reduction in incidence of CPTP would be enough for them to change practice TEB to PVB (or vice-versa). We have powered the trial on a 33% relative reduction which we think is more realistic and likely to be closer to any minimally important difference [14].

### 4.8. Framework

The objective of the trial is to test the superiority of one intervention to another.

The null hypothesis is that there is no difference in the presence of chronic pain between the intervention groups (PVB vs TEB). The alternative hypothesis is that there is a difference between the groups.

#### 4.9. Interim analyses and stopping guidance

Data analyses will be supplied in confidence to an independent Data Monitoring Committee (DMC), which will be asked to give advice on whether the accumulated data from the trial, together with the results from other relevant research, justifies the continuing recruitment of further participants. The DMC will operate in accordance with a trial specific charter based upon the template created by the Damocles Group. The DMC will meet at least annually as agreed by the Committee and documented in the Charter. More frequent meetings may be required for a specific reason (e.g. safety phase) and will be recorded in minutes.

The DMC will be scheduled to meet prior to the recruitment of the first patient, in a joint meeting with the Trial Steering Committee (TSC), one year after the trial opens to recruitment and then annually thereafter until the trial closes to recruitment.

Additional meetings may be called if recruitment is much faster than anticipated and the DMC may, at their discretion, request to meet more frequently or continue to meet following completion of recruitment. An emergency meeting may also be convened if a safety issue is identified. The DMC will report directly to the Trial Management Group (TMG) who will convey the findings of the DMC to the TSC, funders, and/or sponsors as applicable. The DMC may consider recommending the discontinuation of the trial if the recruitment rate or data quality are unacceptable or if any issues are identified which may compromise participant safety. The DMC may recommend stopping the trial early if any interim analyses demonstrate differences between treatments that were deemed to be convincing to the clinical community.

A separate DMC reporting template will be drafted and agreed by the DMC including an agreement on which outcomes will be reported at interim analyses. The statistical methods stated in this SAP will be followed for the outcomes included in the DMC report, where possible.

#### 4.10. Internal Pilot Progression Rules

The trial contains an internal pilot, running for 12 months from the first site being opened to recruitment in which the following criteria have been set as a guide as to the feasibility of continuing to full recruitment:

- Recruiting at least 218 patients
- Opening at least 13 sites to recruitment

- At least 70% of sites which are open to accrual successfully recruiting a patient

#### 4.11. Timing of final analysis

The final analysis for the trial will occur once all participants have completed their scheduled assessments and the corresponding outcome data has been entered onto the trial database and validated as being ready for analysis. This is provided that the trial has not been stopped early for any reason (e.g. DMC advice or funding body request).

#### 4.12. Timing of other analyses

Not applicable.

#### 4.13. Trial comparisons

All references in this document to 'group' refer to paravertebral blockade (PVB) or thoracic epidural blockade (TEB).

### 5. Statistical Principles

#### 5.1. Confidence intervals and p-values

All estimates of differences between groups will be presented with two-sided 95% confidence intervals (CI), unless otherwise stated. For the primary outcome, a p-value will be produced, with statistical significance considered at the 5% level. Results of the secondary outcomes will be treated cautiously given no adjustment for multiple comparisons will be made (see section 5.2).

A p-value from an adjusted model (adjusted as per the primary outcome; see section 9.4) will be presented for the safety data as per section 9.7 where required. We will not adjust for multiplicity as this is counterproductive for considerations of safety [15].

#### 5.2. Adjustments for multiplicity

No correction for multiple testing will be made.

#### 5.3. Analysis populations

All primary analyses (primary and secondary outcomes including safety outcomes) will be analysed using a modified intention-to-treat (MITT) principle. Participants will be followed up and analysed in the intervention group to which they were randomised provided thoracotomy was undertaken, regardless of adherence with the intervention.

In the first instance, this will only include those participants who had undergone a thoracotomy, were alive at six months (the primary outcome time) and had complete outcomes, but missing data sensitivity analysis will explore the impact of missing responses on the primary outcome in those who:

a) Had thoracotomy, were alive but were lost to follow-up or withdrawn;

b) Had thoracotomy, but later died as so did not have opportunity to complete the primary outcome measure

c) A further missing data sensitivity analysis will also explore the impact of those did not have thoracotomy (i.e. will include all randomised).

See section 9.9 for further details.

5.4. Definition of adherence

Adherence to allocated intervention will be recorded on the intervention form. We will define adherence as those who received the allocated intervention (TEB or PVB) only.

5.5. Handling protocol deviations

A protocol deviation is defined as a failure to adhere to the protocol such as errors in applying the inclusion/exclusion criteria, the incorrect intervention being given, incorrect data being collected or measured, follow-up visits outside the visit window or missed follow-up visits.

We will apply the MITT principle and will include all participants as per the MITT population described in section 5.3 in the analysis. This does not include those participants who fail to have a thoracotomy operation; since CPTP cannot develop in these participants and does not make sense to make the post-baseline assessments detailed in the protocol. These participants will be withdrawn from study post baseline assessment. This also does not include those participants who have specifically withdrawn consent for the use of their data in the first instance; however the impact of this will be explored as per other missing responses. See section 9.9 for details on sensitivity analyses.

Three month follow-up questionnaires will be considered valid provided they have been completed prior to the six month assessment time point. Six month follow-up questionnaires will be considered valid provided they have been completed prior to the twelve month assessment time point. Twelve month follow-up questionnaires will be considered valid provided they have been completed before 18 months post randomisation. If the three and six month questionnaires are completed too late to be considered valid for that particular assessment time and subsequent questionnaires have not been returned they will be considered valid for the subsequent time-point (e.g. a six month form completed at thirteen months will not be valid for six month assessment but will be considered valid for the twelve assessment).

In the first instance, any booklets that are not considered valid will be excluded from analysis. These responses will looked at in a sensitivity analysis, see section 9.9 for further details.

#### 5.6. Unblinding

Not applicable, TOPIC 2 is an open (unblinded) study.

### 6. Trial population

#### 6.1. Recruitment

A flow diagram (as recommended by CONSORT [16]) will be produced to describe the participant flow through each stage of the trial. This will include information on the number who proceeded to thoracotomy following randomisation, alongside withdrawals (dropouts and losses to follow up) and death before reaching the assessment times. A template for reporting this is given in Appendix D1.

#### 6.2. Baseline characteristics

The trial population will be tabulated as per Appendix D2. Categorical data will be summarised by number of participants, counts and percentages. Continuous data will be summarised by the number of participants, mean and standard deviation if deemed to be normally distributed or number of participants, median and interquartile range if data are skewed, and ranges if appropriate. Tests of statistical significance will not be undertaken, nor confidence intervals presented [17].

### 7. Intervention(s)

#### 7.1. Description of the intervention(s)

A template for reporting information on the intervention(s) is given in Appendix D3.

#### 7.2. Adherence to allocated intervention

A cross-tabulation of allocated intervention by the adherence categories stated in section 5.4 will be produced (proportions and percentages). A template for reporting adherence is given in Appendix D8.

### 8. Protocol deviations

Frequencies and percentages by group will be tabulated for the protocol deviations as per Appendix D9.

### 9. Analysis methods

Intervention groups will be compared using regression models appropriate for the data type to adjust for all covariates as specified in section 9.1, where possible.

#### 9.1. Covariate adjustment

In the first instance, intervention effects between groups for all outcomes will be adjusted for the minimisation parameters listed in section 4.6. These parameters will be included in the model as categorical data. All variables in the model will be treated as fixed effects apart from centre which will be included as a random effect. Other covariate adjustment will be baseline values for parameters, where appropriate (e.g. include the baseline score as a covariate in the model).

#### 9.2. Distributional assumptions and outlying responses

Distributional assumptions (e.g. normality of regression residuals for continuous outcomes) will be assessed visually prior to analysis; although in the first instance the proposed primary method of estimation in this analysis plan will be followed. If responses are considered to be particularly skewed and/or distributional assumptions violated, other analytical options will be considered such as the transformation of responses prior to analysis (e.g. log transformation) or the use of medians and interquartile ranges alongside unadjusted differences in medians using bootstrapping methods.

If extreme values are apparent and considered to be affecting the integrity of the analysis, a sensitivity analysis consisting of removing the outlying response(s) and repeating the analysis will be performed. Output from these analyses, if performed, will be described and presented alongside the original analysis (or included, e.g. in appendices) with the excluded values clearly labelled. See section 9.9 for further details regarding sensitivity analyses.

#### 9.3. Handling missing data

In the first instance, analysis will be completed on received data only with every effort made to follow-up participants to minimise any potential for bias. To examine the possible impact of missing data on the results, sensitivity analysis will be performed on the primary outcome measure [18]. See section 9.9 for further details regarding sensitivity analyses.

#### 9.4. Analysis methods – primary outcome (s)

A template for reporting the primary outcome is given in Appendix D11. Summary statistics (numbers and percentages) will be presented by intervention group.

A generalised estimating equations (GEE) model will be used to calculate the risk difference (identity link) and relative risk (log link) with 95% confidence intervals for the primary outcome (Worst pain) taking into account all timepoints in the chronic phase and adjusting for the variables listed in section 9.1. An independent covariance structure will be assumed. Time will be assumed to be categorical (fixed) variable. To account for a temporal effect, a time by treatment interaction parameter will be included in the model (estimates of the differences between groups at relevant time be taken from the model including this interaction parameter).

If the model fails to converge then, in the first instance, a Poisson regression model with randomising centre as a random effect and robust standard errors will be used to estimate the same parameters [19]. In the case that the Poisson regression model still does not converge, randomising centre will be omitted, and the log-binomial model will be refit for just fixed effects. If this also fails, other covariates that are not the treatment group will be dropped. A Poisson model will not estimate the risk difference, so if the initial model fails to converge, the same process of variable removal will be followed, within a log-binomial model framework. The p-value relating to the intervention group parameter as generated by the model estimating the relative risk will be presented. These type of models will not take into account follow-up at all time-points; they will be utilised separately for each follow-up time (time and time by treatment interaction will not be included).

See section 9.1 for further details on covariate adjustment.

#### 9.5. Analysis methods – secondary outcomes

A template for reporting the secondary outcomes is given in Appendix D13.

Questionnaire scores:

1. Continuous outcomes: Acute pain in the 3 days following surgery and at discharge, completed by the patient (via VAS [at each day and the total over all 3 days]; BPI)

2. Continuous outcome: SF-MPQ-2 at discharge

3. Continuous outcomes: Chronic pain (via VAS, BPI and SF-MPQ-2, completed by the participant at 3-, 6- and 12-months post randomisation)

4. For dichotomous secondary outcomes listed below: relative risks, risk differences and confidence intervals will be generated in the same fashion as the primary outcome.

- Presence of severe pain at its worst (VAS>=70) in chronic phase at 3, 6 and 12 months
- Presence of moderate pain at its worst (VAS>=40) in chronic phase at 3 and 12 months
- Presence of severe pain on average (VAS>=70) in chronic phase at 3, 6 and 12 months
- Presence of moderate pain on average (VAS>=40) in chronic phase at 3, 6 and 12 months

5. Continuous outcomes: General health-related quality of life (by EQ-5D-5L, completed by the participant at discharge and at 3, 6 and 12 months) and Mental health state (measured by HADS, completed by the participant at discharge and at 3, 6 and 12 months.

6. Ordered categorical outcomes: Patient satisfaction (by Likert scale), completed by the participant at discharge and at 3, 6 and 12 months; severe pain responses in chronic phase at 3, 6 and 12 months (0-9 'no pain'; 10-39 'mild pain'; 40-69 'moderate pain'; 70-100 'severe pain'); moderate pain responses in chronic phase at 3, 6 and 12 months (0-9 'no pain'; 10-39 'mild pain'; 40-69 'moderate pain'; 70-100 'severe pain')

For points 1, 3 and 5 above: Difference between group means and associated confidence intervals at each time point will be estimated using a mixed repeated measures linear regression model, including all assessment times (fixed effect). Parameters allowing for treatment group, time and the minimisation variables (section 4.6) will be included (all as fixed effects). Recruiting centre will be included in the model as a random effect. Time will be assumed to be categorical (fixed) variable. To account for a temporal effect, a time by treatment interaction parameter will be included in the model (estimates of the differences between groups at relevant time be taken from the model including this interaction parameter). A general ‘unstructured’ covariance structure will be assumed. In the case that the distribution of continuous outcomes are skewed this data is best summarised by medians and interquartile ranges, unadjusted median differences will be estimated at each time point; confidence intervals will be obtained by utilising bootstrapping methods.

Furthermore, for point 5 only, the models will also include baseline variables.

For point 2, SF-MPQ-2 at discharge will be analysed using linear regression to obtain mean differences between groups and 95% confidence intervals. Minimisation variables will be included in the model.

With respect to point 6, a GEE model with alternating logistic regression will used taking into account each timepoint. A common odds ratio and 95% confidence intervals will be generated. A fully exchangeable working correlation structure will be used. Time will be assumed to be categorical (fixed) variable. To account for a temporal effect, a time by treatment interaction parameter will be included in the model (estimates of the differences between groups at relevant time be taken from the model including this interaction parameter).

Analgesic use – this will be standardised to Morphine levels and summarised using summary statistics (mean (SD) or median [IQR] as appropriate).

#### 9.6. Analysis methods – exploratory outcomes and analyses

Not Applicable.

#### 9.7. Safety data

With the following points below, number and percentages of participants will be presented by intervention group:

- Complications of regional analgesia (failure of blockade, hypotension (systolic blood pressure (<90mmHg)), inadequate pain relief, low respiratory rate (<10/minute), drowsiness, nausea and vomiting, urinary retention, itching, high block, post-dural puncture headache, vascular puncture, pleural puncture).

- Occurrence and severity of surgical complications until discharge from hospital (occurrence as defined by the European Society of Thoracic Surgeons dataset and severity as defined by the TMM classification, including any occurrence
- Post-operative pulmonary complications (PPCs) until discharge from hospital (as defined by SteP COMPAC), including any occurrence
- Critical care admission (levels 2 and 3)
- Mortality (reported for all deaths due to all causes)
- Serious Adverse Events
- Suspected unexpected serious adverse reactions (SUSARs)

Statistical significance will be determined by chi-squared test for SAEs and SUSARs combined. If the rate of any of the other safety outcomes listed above exceeds 2% we will consider formal analysis in a similar fashion to SAE/SUSAR outcome. A template for reporting this safety data is given in Appendix D14.

### 9.8. Planned subgroup analyses

Interpretation of subgroup analysis will be treated with caution (output will be treated as exploratory rather than definitive [20]). Analysis will be limited to the primary outcome (at 6 months only), and the following subgroups:

- Gender
- Age (<65 years, ≥65 years)
- Thoracotomy for lung cancer resection or for other indication

The effects of these subgroups will be examined by adding the subgroup by treatment group interaction parameters into the model. Statistical significance (p-values) of these interaction parameters will be determined by Wald tests. Differences between the treatment groups within subgroups will only be examined if the interaction parameter is shown to statistically important (p-value <0.05)

A template for reporting the subgroup analyses for the primary outcome is given in Appendix D15.

### 9.9. Sensitivity and supportive analyses

Sensitivity analyses will be limited to the primary outcome and will consist of:

- Where all returned forms regardless of whether they were returned in the correct time window are included.
- Where a ‘CPTP’ event is defined to be a VAS score greater than or equal to 40 at both 3 and 6 months (worst pain). This is predominantly to assess whether the participants have experienced pain for at least 3 months.
- A per-protocol analysis following the methods described in section 9.4 including only participants who received the intervention that they were randomised to and did not receive an alternative intervention at any point if the block was re-sited.
- A ‘tipping point’ analysis will be carried out for the participants who have missing primary outcome data. This analysis will explore the possibility that these missing responses are ‘missing not at random’ (MNAR) using a tipping point approach. The general methodology is described below but will be conducted in a number of different populations: a) in those who had thoracotomy, were alive but had missing follow-up; b) had thoracotomy, but later died as so did not have opportunity to complete the primary outcome measure as well as population a); and c) all randomised; so missing are those who did not undergo thoracotomy as well as populations a) and b). For those who were missing due to not having undergone surgery or due to death, the assumption here is these events are not related to the probability of being missing (i.e. the intervention would have no impact on these events). In any tipping point analysis, the initial rate applied will be equal to the observed rate in the both of the groups.
- The method that will be described in each population is as follows:

o In this analysis, for participants with missing outcome data, events will be added sequentially in each of the groups in turn to determine the point where the general conclusion changes (for example from positive to inconclusive in terms of the CI). An assessment can then be made about whether the event rate in the missing responses between groups is likely to be plausible when compared with the event rate in the non-missing data. Two scenarios will be considered as follows:

o Scenario A: In the usual care arm (TEB), assume all missing responses are non-events (i.e. No Pain). For missing responses in the PVB group, we will first replace X missing responses with events (i.e. Pain) where X is the number of events such that the event rate in the missing responses is equal to the event rate in the non-missing responses in the PVB group (for example if the event rate in the non-missing data in the PVB group is 8% and we have 20 missing responses we will add 0.08*20 ~ 2 events [number of events rounded to the closest integer >0]). This will be regarded as the base case. All other missing responses in the PVB group will be considered as non-events. An unadjusted model will be fitted. Event rates will be compared between groups. The CI from the treatment estimate (from the RR of the log-binomial model) will be examined and stored. Following the base case, an additional event will be added to the PVB group and the above procedure repeated (unadjusted model fitted and the RR and CI examined). This process will end when the number of events added to the PVB group are equal to the original number of missing responses in this group. The tipping point for the PVB group will occur when enough events have been added such that the upper/lower limit of the CI from the corresponding model differs from that of the primary ITT finding (in regards to whether the CI crosses the null value of one).

o Scenario B: In the PVB group, assume all missing responses are non-events (i.e. No Pain). For missing responses in the TEB group, we will first replace X missing responses with events (i.e. X additional cases with Pain) where X is the number of events such that the event rate in the missing responses is equal to the event rate in the non-missing responses in the TEB group (for example if the event rate in the non-missing data in TEB group is 15% and we have 20 missing responses we will add 0.15*20 ~ 3 events [number of events rounded to the closest integer >0]). This will be regarded as the base case. All other missing responses in the TEB group will be considered as non-events. An unadjusted model will be fitted. Event rates will be compared between groups. The CI from the treatment estimate (from the RR of the log-binomial model) will be examined and stored. Following the base case, an additional event will be added to the TEB group and the above procedure repeated (unadjusted model fitted and the RR and CI examined). This process will end when the number of events added to the placebo group are equal to the original number of missing responses in this group. The tipping point for the TEB group will occur when enough events have been added such that the upper/lower limit of the CI from the corresponding model differs from that of the primary ITT finding (in regards to whether the CI crosses the null value of one).

o For both scenario A and scenario B, results will be presented visually. The event rate (CPTP incidence rate) in the missing data in the PVB group (scenario A) or TEB group (scenario B) will be plotted on the x-axis. The RR (PVB vs. TEB) from the model will be on the plotted on the y-axis. The corresponding CI will be included around each RR. If the CI in the ITT analysis contains one, then the point where the upper CI falls below or above one will be highlighted (tipping point) on the plot. If the CI in the ITT analysis does not contain one (i.e. one treatment is superior to the other), then the point where the CI crosses one will be highlighted (tipping point). The base case will also be highlighted on each plot.

### 10. Analysis of sub-randomisations

Not Applicable.

### 11. Health economic analysis

As indicated in the protocol there will also be an economic analysis. The details of this analysis are documented separately.

12. Statistical software

Statistical analysis will be undertaken in the following statistical software packages: SAS software, version 9.4 (or higher) or STATA version 17 (or higher)
